# A Call For Better Methodological Quality Of Reviews On Using Artificial Intelligence For COVID-19 Detection In Medical Imaging – An Umbrella Systematic Review

**DOI:** 10.1101/2021.05.03.21256565

**Authors:** Paweł Jemioło, Dawid Storman, Patryk Orzechowski

## Abstract

**Objective:** In this umbrella systematic review, we screen existing reviews on using artificial intelligence (AI) techniques to diagnose COVID-19 in patients of any age and sex (both hospitalised and ambulatory) using medical images and assess their methodological quality.

**Methods:** We searched seven databases (MEDLINE, EMBASE, Web of Science, Scopus, dblp, Cochrane Library, IEEE Xplore) and two preprint services (arXiv, OSF Preprints) up to September 1, 2020. Eligible studies were identified as reviews or surveys where any metric of classification of detection of COVID-19 using AI was provided. Two independent reviewers did all steps of identification of records (titles and abstracts screening, full texts assessment, essential data extraction, and quality assessment). Any discrepancies were resolved by discussion. We qualitatively analyse methodological credibility of the reviews using AMSTAR 2 and evaluate reporting using PRISMA-DTA tools, leaving quantitative analysis for further publications.

**Results:** We included 22 reviews out of 725 records covering 165 primary studies. This review covers 416,254 participants in total, including 50,022 diagnosed with COVID-19. The methodological quality of all eligible studies was rated as critically low. 91% of papers had significant flaws in reporting quality. More than half of the reviews did not comment on the results of previously published reviews at all. Almost three fourth of the studies included less than 10% of available studies.

**Discussion:** In this umbrella review, we focus on the descriptive summary of included papers. Much wasting time and resources could be avoided if referring to previous reviews and following methodological guidelines. Due to the low credibility of evidence and flawed reporting, any recommendation about automated COVID-19 clinical diagnosis from medical images using AI at this point cannot be provided.

**Funding:** PO was supported by NIH grant AI116794 (the funding body had no role in the design, in any stage of the review, or in writing the manuscript); PJ and DS did not receive any funding.

**Registration:** The protocol of this review was registered on the OSF platform [1].

## 1 Introduction

In early December 2019, a new coronavirus epidemic was identified in Wuhan, Hubei Province, China [2]. Coronavirus disease 2019 (COVID-19) is a viral infection spread by direct contact with people with the illness (from droplets generated by sneezing and coughing), or indirectly [3]. It is caused by Severe Acute Respiratory Syndrome Coronavirus 2 (SARS-CoV-2). As of May 3, 2021, over 152 million people have been diagnosed with COVID-19, with nearly 3.2 million associated deaths^1^. Two first waves of COVID-19 affected many societies, as well as scientific organisations [4–7]. On January 30, 2020, the World Health Organisation (WHO) issued a public health emergency of international concern (PHEIC) associated with COVID-19, and declared the state of a pandemic on March 11, 2020 [8].

Disease manifestation is variable, with some infected people remaining asymptomatic (even up to 10% [9]) and others suffering from mild (including fever, cough and aches) to severe (involving lethargy with dyspnea and increased respiratory rate) and critical manifestations (requiring mechanical ventilation). It may lead to serious neurological, musculoskeletal, or cerebrovascular disorders or may even progress to a life-threatening respiratory syndrome in some patients [10, 11]. Moreover, in 80% of patients, COVID-19 leaves one or more long-lasting symptoms, with fatigue, headaches, attentional difficulties, anosmia, memory loss manifested the most frequently [12]. Wide-ranging longer-term morbidity has also been described in the absence of a severe initial illness [13].

With limited access to healthcare and qualified personnel due to pandemic, an early diagnosis of COVID-19 is essential in preventing transmission of the virus. Identification of infected allows for better management of the pandemic (e.g. isolation, quarantine, hospital admission or admission to the intensive care unit) [14]. Understanding the accuracy of tests and diagnostic features seems essential to develop effective screening and management methods [15].

There are many diagnostic evaluation challenges due to the COVID-19 pandemic. The primary method for diagnosing COVID-19 is Nucleic Acid Amplification Tests (NAATs) that use different methods to amplify nucleic acids and detect the virus, e.g. strand displacement amplification, reverse transcription-polymerase chain reaction (RT-PCR), or transcription-mediated amplification [16]. It utilises respiratory tract samples (mainly from the nasopharynx or oropharynx). However, some guidelines recommend nasal swabs [16], and some evidence suggests lower respiratory samples, such as sputum, may have higher sensitivity [17]. Serological tests are being used for detecting antibodies to SARS-CoV-2 for confirmation of past infection [16]. Nevertheless, there are concerns regarding the evidence for their accuracy and value in specific populations and clinical situations [18]. Additionally, rapid antigen and molecular-based tests are also available, but their diagnostic effectiveness is still unclear [16].

At the same time, the role of medical imaging in COVID-19 patients is emerging. It provides fast and sound insight into the lungs status and thus is used in emergency departments [15]. From the pandemic onset, chest radiography (X-Ray) has been used as a helpful tool for COVID-19 diagnosis [19]. However, diagnosing based on X-Ray is challenging, as even a normal chest radiography does not confirm that the patient has COVID-19, especially when it is an early stage [20].

A computed tomography (CT) has been able to discover COVID-19 abnormalities with sensitivity exceeding 97%. Some evidence suggests it helps to detect COVID-19 earlier than manifested by the positive RT-PCR test [21, 22]. CT as a diagnostic tool for COVID-19 was reported to have over 90% sensitivity, but only from 25% to 83% of specificity for symptomatic patients [23]. Positive COVID-19 diagnosis using CT has not been sufficient to determine the severity and the outcome of the disease [21].

With the increasing role of medical imaging as a diagnostic tool for COVID-19, a question arises if and to which extent automated tools can be included in clinical diagnosis. Up to this day, artificial intelligence (AI), or more specifically, deep learning (DL), have started to play an increasingly vital role in medicine [24]. In some recent studies and clinical trials, AI has been demonstrated to match or even exceed the performance of expert radiologists, which could potentially offer expedited and less expensive diagnostics [25–30]. A recent study and meta-analysis by Li et al. [31] with 31,587 identified and 82 included studies shows deep learning is capable of slightly outperforming health care professionals in detecting diseases from medical images with a pooled sensitivity of 87% (vs 86% of health care professionals) and pooled specificity of 93% (vs 91% respectively). Overlapping confidence intervals suggest that there is no statistically significant difference in performance between AI and human.

Since the onset of the pandemic, multiple initiatives have been taken in order to share data and knowledge on COVID-19 between researchers [32], just to mention COVID-19 Open Research Dataset Challenge (CORD-19) database with over 400,000 articles (over 150,000 with full text) as of May 3, 2021 on COVID-19 and related diseases ready for data mining on a popular Kaggle server for data science^2^ or Global literature on coronavirus disease^3^ led by World Health Organisation.

The surge of primary studies is followed by the rise of the reviews summarising previous work. However, the emergency of the COVID-19 topic seemed to have led to increase overlap and degradation of the quality of the research [33].

This umbrella systematic review aims to screen existing reviews on using artificial intelligence (AI) techniques to diagnose COVID-19 in patients of any age and sex (both hospitalised and ambulatory) using medical images and assess their methodological quality. We provide evidence that reliable systematic reviews regarding this topic are in great need. The extended quantitative analyses, as well as in-depth credibility exploration, will be published separately.

## 2 Methods

### 2.1 Eligibility criteria, Protocol

We focused on any review (systematic or not) that includes primary studies utilising AI methods with medical imaging results to diagnose COVID-19. We were particularly interested in the performance of such classification systems, e.g. accuracy, sensitivity, specificity. We excluded these primary studies that used reference standards other than assay types (NAATs, antigen tests, and antibody tests) from nasopharyngeal or oropharyngeal swab samples, nasal aspirate, nasal wash or saliva, sputum or tracheal aspirate, or bronchoalveolar lavage (BAL) [16, 34]. Additionally, due to overlapping and double referencing of the post-conferencing articles (particular chapters), we excluded entire proceeding and post-conference books as they contain little information about the topics (presented in chapters) *per se*. However, we did not exclude reviews (chapters) as they were still present in our search. The protocol of this review was published [35] and registered [1] on the OSF platform.

### 2.2 Search methods

In order to determine whether there are any eligible papers, we conducted a pre-search in the middle of August 2020 via Google Scholar by browsing. We searched seven article databases (MEDLINE, EMBASE, Web of Science, Scopus, dblp, Cochrane Library, IEEE Xplore) and two preprint databases (arXiv, OSF Preprints) from inception to 01 September 2020 using predefined search strategies. We checked included preprints for peer-reviewed version before extraction phase.

In developing the search strategy for MEDLINE, we combined the Medical Subject Headings (MeSH) and full-text words. They concerned artificial intelligence, diagnostics, imaging and COVID-19. We adapted the MEDLINE strategy for other sources searched. In Appendix A, we present used strategies. No date or language restrictions were adopted. Additionally, we searched references of included studies for references.

### 2.3 Definitions

We defined the terms used in our eligibility criteria below. *Review* refers to a paper identified by authors as a review or a survey. *AI* refers to computer programs that can perform tasks as intelligent beings [36]. *COVID-19* refers to a disease caused by the SARS-CoV-2 virus [37]. *Imaging* refers to individuals medical imaging results (e.g., CT scans, X-rays, ultrasound images) [38, 39]. *Diagnosis* refers to the identification of an illness (here: COVID-19)^4^. *Performance metrics* refers to evaluating machine learning algorithms. These measures are utilised to juxtapose observed data (actual labels) with the predictions of the model [40].

### 2.4 Data collection

Using Endnote (Claritive Analytics ®) and Rayyan [41], we checked identified references for duplicates. PJ, DS, PO independently screened the remaining references using the latter application. Subsequently, the reviewers (PJ, DS, PO) assessed the full texts for meeting the inclusion criteria separately. We pre-specified an extraction form, and PJ and DS collected all necessary data independently. As specified in the protocol [1], we gathered information about authors, funding, population, models, outcomes – AI methods performance, and additional analyses, e.g. interpretability.

To improve the understanding of the criteria among the reviewers, we carried out pilot exercises before each phase of this umbrella review (screening of titles and abstracts, full texts assessment, data extraction, credibility assessment). We achieved consensus via discussion if any conflicts (at each step of identification) occurred.

### 2.5 Quality assessment

The evaluation was conducted independently by PJ and DS. All discrepancies were resolved by discussion. We assessed the methodological credibility using an extended version of the QASR tool [42] and AMSTAR 2 [43] with critical items (2, 4, 7, 9, 11, 13, and 15). In this study, we present only results from the latter instrument since the comprehensive assessment of included reviews will be published separately. The general quality across the study was assessed as critically low when at least one item in a critical domain was evaluated as a flaw.

Additionally, we assessed the quality of reporting in included studies using the PRISMA-DTA Checklist [44]. We rated each module on the 3-item scale: 0 (no with no compliance), 0.5 (partial yes with fragmentary compliance), 1 (yes with total compliance). Next, the results were summed, and the overall score was then assigned. Applying the Li et al. method [45], we differentiated the quality of reporting as follows:

- major flaws when the final score was ≤ 15.0,
- minor flaws when the final score was between 15.5 and 21.0,
- minimal flaws when the final score was ≥ 21.5.

### 2.6 Analyses

In this umbrella review, we focus on the descriptive summary of included papers regarding quality and reporting on population, models, and interpretability. To identify a scale of wasting of time and resources, we also extracted bibliometric data about publishing dates (availability), sending to the editors (first and last version), and acceptance. Moreover, we checked how much evidence overlap across included reviews. In all tables and figures, included reviews are sorted by date of the last received version of the article; if it was not available, we used acceptance or published date (available online), whichever first occurred.

The quantitative analysis and following additional analyses will be published separately, as well as extended credibility evaluation. Thus, we do not present a subgroup analysis and investigation of heterogeneity or sensitivity analysis in this paper.

## 3 Results

### 3.1 Included studies

After removing duplicates, we screened 725 studies, of which 33 were read in the form of full texts. In total, we included 22 reviews [46–67] for qualitative synthesis. Our reporting is consistent with Preferred Reporting Items for Systematic Reviews and Meta-analyses (PRISMA) guidelines [68]. The full study flow is presented in Figure 1.

**Figure 1:**
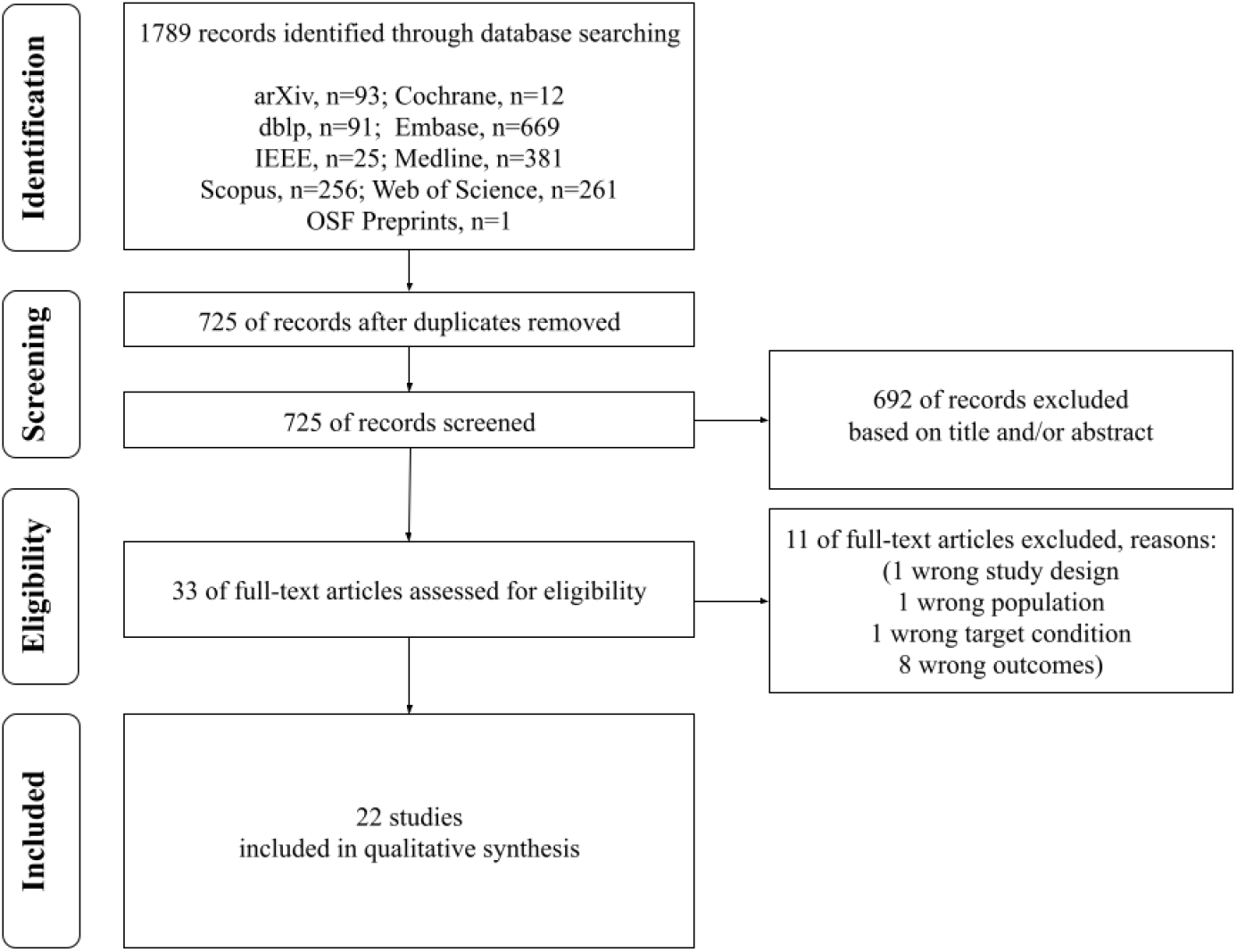
PRISMA flow chart.

The bibliometric characteristics of included studies are shown in Table 1 and in Appendix D. The lists of included and excluded studies (with reasons) are presented in Appendices B and C, respectively. None of the studies included meta-analysis, but one summarised the results as an average without weights [60].

**Table 1:** Basic characteristics of included studies.

| Variable | N (%) | Mean (range) |
| --- | --- | --- |
| Country |  |  |
| USA | 8 (18%) | NA |
| Australia | 4 (9%) | NA |
| China | 4 (9%) | NA |
| India | 4 (9%) | NA |
| UK | 3 (7%) | NA |
| other | 22 (49%) | NA |
| Number of authors | 171 | 8 (1-43) |
| Source |  |  |
| Preprint | 8 (36%) | NA |
| IEEE Access | 2 (9%) | NA |
| IEEE Reviews in Biomedical Engineering | 2 (9%) | NA |
| Diagnostic and Interventional Imaging | 2 (9%) | NA |
| Diabetes & Metabolic Syndrome:<br>Clinical Research & Reviews | 1 (5%) | NA |
| Applied Intelligence | 1 (5%) | NA |
| British Medical Journal | 1 (5%) | NA |
| Biosensors and Bioelectronics | 1 (5%) | NA |
| Machine Vision and Applications | 1 (5%) | NA |
| Current Problems in Diagnostic Radiology | 1 (5%) | NA |
| Journal of the Indian Medical Association | 1 (5%) | NA |
| International Conference on Nanotechnology | 1 (5%) | NA |
| Impact Factor | NA | 4.1 (0-30.313) |
| Is this a Systematic Review? | 2 (9%) | NA |
| Sources searched | 50 | 5 (3-7) |
| arXiv | 8 (36%) | NA |
| medRxiv | 6 (27%) | NA |
| Pubmed/Medline | 6 (27%) | NA |
| Google Scholar | 6 (27%) | NA |
| bioRxiv | 5 (23%) | NA |
| IEEE Xplore | 3 (14%) | NA |
| Science Direct | 3 (14%) | NA |
| ACM digital library | 2 (9%) | NA |
| Springer | 2 (9%) | NA |
| MICCAI conference | 1 (5%) | NA |
| IPMI conference | 1 (5%) | NA |
| Embase | 1 (5%) | NA |
| Web of Science | 1 (5%) | NA |
| Elsevier | 1 (5%) | NA |
| Nature | 1 (5%) | NA |
| Number of studies |  |  |
| included | 358 | 51 (20-107) |
| interesting | 451 | 21 (1-106) |
| observational | 26 (6%) | NA |
| unclear study design | 425 (94%) | NA |

There were 399,374 participants in total in analysed studies. 46,753 of them suffered from COVID-19 (12%). None of the reviews provided information about ethnicity, smoking, and comorbidities (diabetes mellitus, cardiovascular disorders, obesity, respiratory diseases, autoimmunological, or others). Only one review [57] reported on age and gender proportion. Table 2 presents detailed features.

**Table 2:** Additional characteristics of included studies.

| Variable | N (%) | Mean (range) |
| --- | --- | --- |
| Number of participants |  |  |
| total | 399,374 | 2,256 (25-32,717) |
| COVID-19 | 46,753 | 320 (25-3,389) |
| control | 212,366 | 1,646 (25-23,664) |
| Age |  |  |
| Reported | 1 (5%) | NA |
| Not Reported | 21 (95%) | NA |
| Number of females |  |  |
| Reported | 1 (5%) | NA |
| Not Reported | 21 (95%) | NA |
| Was architecture discussed? | 15 (68%) | NA |
| Was interpretability discussed? | 2 (9%) | NA |
| Number of different experiments with models | 528 | 24 (1-115) |

### 3.2 Quality of included studies

The general quality of all included studies is critically low (see Figure 2). 6 [51–53,55,57,66] reviews provided full information about sources of funding and conflict of interest. It was the most satisfied item. None of the studies provided a list of excluded papers, explanation of eligible study design and sources of funding in included studies.

**Figure 2:**
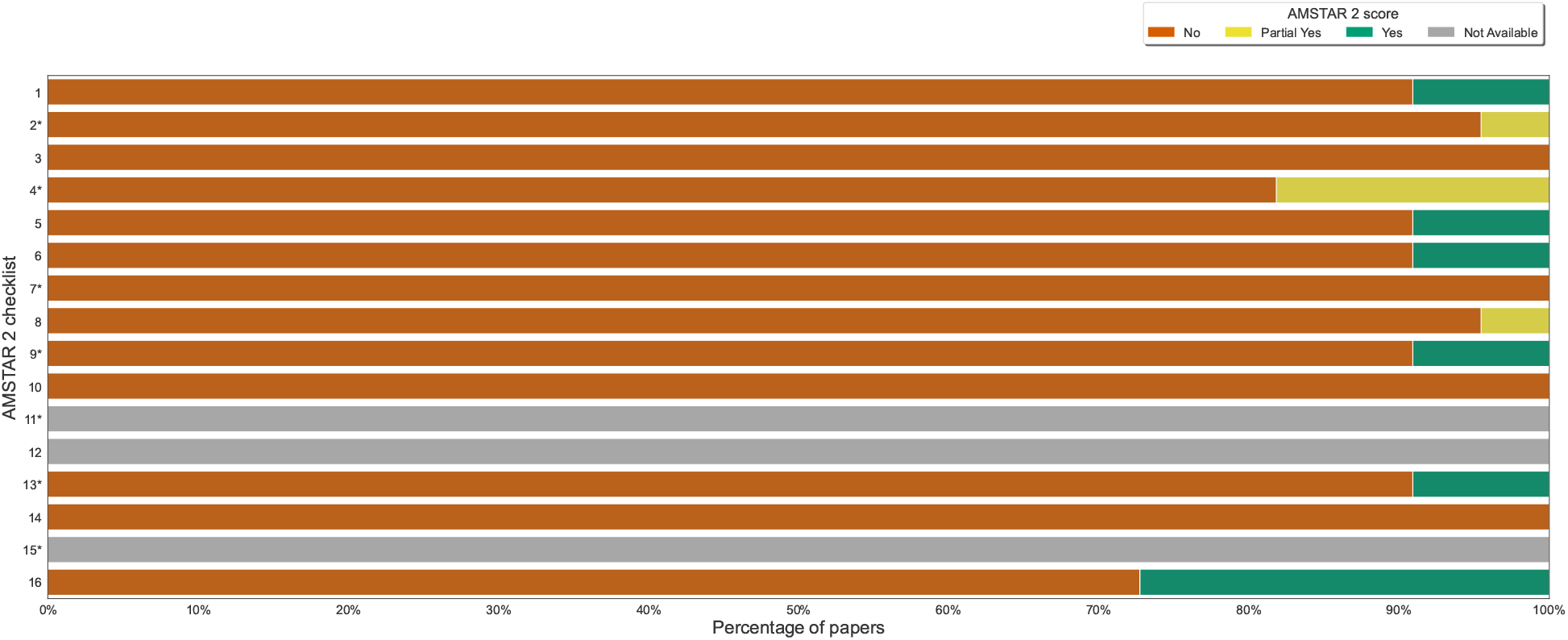
Quality graph: our judgements on each AMSTAR 2 item presented as the percentage of all included studies.

A heatmap with all authors’ judgements regarding AMMSTAR 2 items per specific review can be found in Appendix H. In Appendix I, we also included summarised results per specific review.

Due to the lack of quantitative synthesis in the included reviews, we decided to lower the cut-offs for the overall reporting quality assessment. The thresholds were changed as follows: ≤14.5, 14.5-21.0, and ≥21.0 for major, minor or minimal flaws, respectively.

Major flaws are present among 20 of included papers. All the rest [57,66] reviews contain minor drawbacks (see Figure 3 and Appendices F and G). The most affected domains were these concerning additional analyses both in terms of methods and results (all reviews). Similarly, a summary of evidence was not reported in any review.

**Figure 3:**
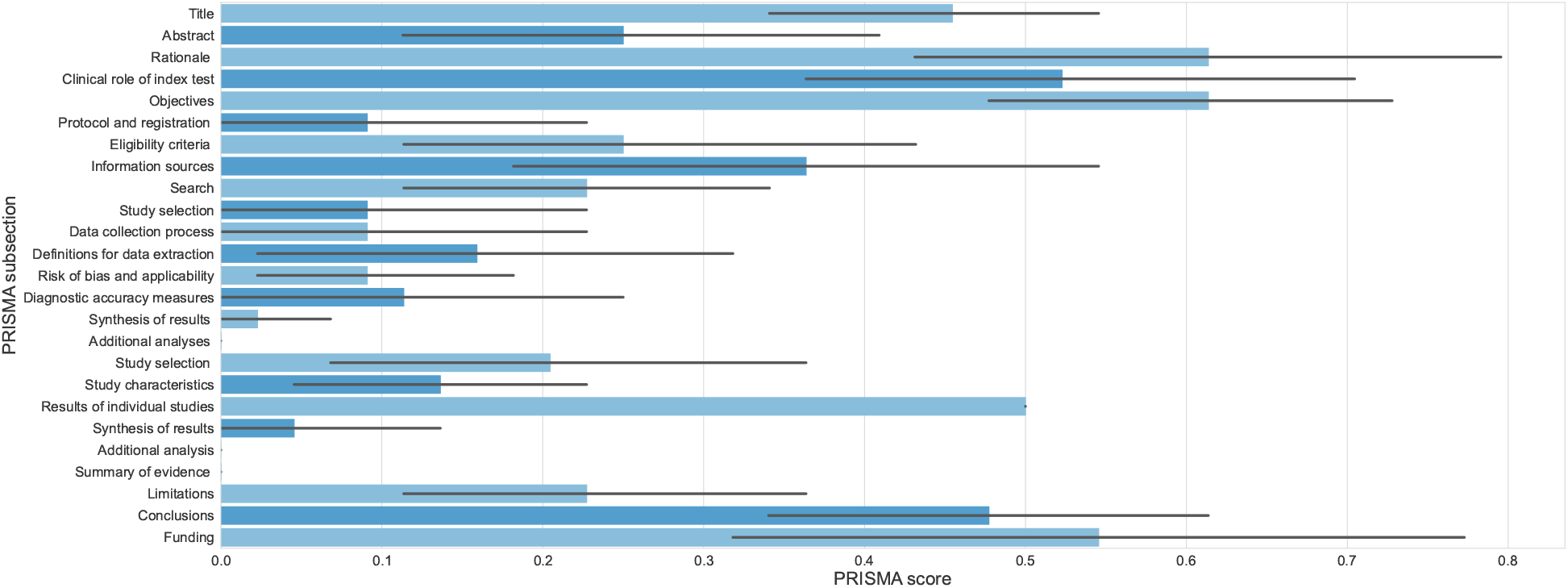
Quality of reporting graph: our judgements about each PRISMA-DTA item presented as averages (with 95% confidence intervals) across all included studies.

Furthermore, 12 of included papers ([46, 48, 51–53–55–57–60–66–67]) reported fully on funding. Additionally, 11 reviews ([46, 47, 49, 54, 55, 58, 59, 61, 62, 65, 66]) and 8 reviews ([46, 48, 49, 54–56–60–66]) described rationale and objectives in the introduction adequately. These were the most satisfied domains.

The mean overall score of reporting quality across the included reviews equals 6.23 (1.5-17.5). Across all items and studies, the most frequent score was 0 (*no*) with 67%. 19% of the times, we assessed the items as 0.5 (*partially yes*).

A heatmap with all authors’ judgements regarding PRISMA-DTA items per specific review can be found in Appendix F. In Appendix G, we also included summarised results per specific review.

### 3.3 Analyses

The included studies were published or available online without peer-reviewing from April 11, 2020 to October 12 2020 (see Appendix B). In Figure 4, we presented a cumulative (from April to September 2020) chart of all 165 primary studies included in discussed reviews. The number of included interesting studies (related to our research question) in selected reviews ranged from 1 [51–53] to 106 [62]. Additionally, we present the percentage of unique articles per review. Unique means that such a study was introduced by a particular review.

**Figure 4:**
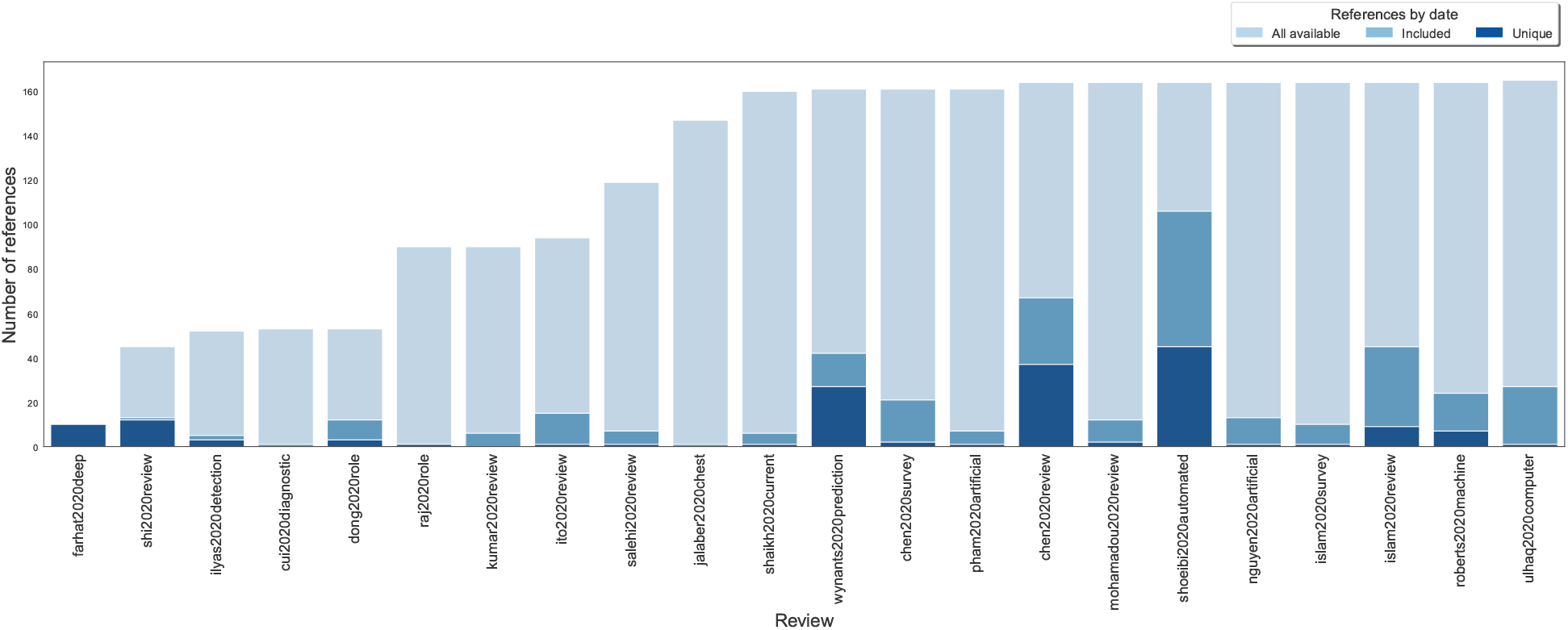
The cumulative chart of included, available, and unique primary papers among included reviews.

50% of all primary studies (the half-saturation constant) were included at least once before the end of July. However, the same number of papers was available for inclusion three months earlier.

Investigating the citations between the reviews, we considered two different scenarios: citing only published reviews and citing both published and preprint versions. In the first case, 0.81 (0-4) reviews were cross-cited. While in the second condition, 1.1 (0-7) published papers or preprints were quoted by the authors of the subsequent reviews. Notably, 12 (55%) of reviews did not cite any previous reviews at all.

In the next step, we analysed how extensive was the search performed by the authors of the reviews, i.e. to which extent the authors identified the primary studies available up to reference date. By a reference date, we considered the day that the review was either received, or accepted by the editors, or published (See Appendix B). The mean percentage of the primary studies covered was 14%, (1-64). Relaxing this condition to the date the last cited paper included in the review was available, the mean percentage of covered studies increased to 24% (1-65). For the full details, see Table 10 (Appendix E).

14% of studies did not include any new paper into consideration. The mean unique primary studies included in particular review was 7.24 (0-45).

## 4 Discussion

Since the emergence of COVID-19 pandemic, over 120,000 related papers (and growing) were published, according to the LitCovid tracking website [69] and NIH OPA iSearch COVID-19 Portfolio^5^. The urgency of reporting novel findings and high pressure to publish COVID-19 related research quickly has been reported to lead to exceptions to high standards of quality [70, 71], lowering methodological credibility of some of the articles [72], or even accepting papers with many analytical errors [73]. Even the quality of the papers published in the three of the most prestigious journals in the field of healthcare: the New England Journal of Medicine, The Lancet, and the Journal of the American Medical Association, was reported to be below their average [74]. Lowered methodological credibility of COVID-19 research papers was also observed in 686 COVID-19 screened articles extracted by Jung et al. from 14,787 COVID-19 papers [75]. An analysis of reviews on COVID-19 by Yu et al. [76] also showed their unsatisfactory credibility. Out of 17 available reviews published before September 1, 2020, 5 (29%) were found to be of low and the remaining 12 (71%) of critically low quality. This is also in line with Li et al. [31] who assessed 63 systematic reviews (25%) to have low and 150 (62%) to have critically low quality. The authors also evaluated reporting using PRISMA [77], and the median score was 14 (10-18). This adds on top of generally low quality of reporting of DL performance from medical images, with high risk of bias present in 58 out of 81 of the existing studies (72%) [78].

Similar conclusions come from our study – we report that the quality of the included reviews was critically low. There were several critical domains that all the reviews failed to meet (selection criteria, list of excluded studies, discussing heterogeneity). Poor quality is not related only to COVID-19 articles and AI. Still, it occurs in many fields like bariatric surgery with 99% critically low articles [79], psychology with 95% [80] or methodological where 53 out of 63 publications were critically low of quality [81]. What is more, the reporting was also poor. Three PRISMA-DTA domains were fully flawed in all reviews (additional analyses in methods and results descriptions, summary of evidence and discussion). Only in one review [64], the authors provided description of methods used for synthesis partially. Although the included studies focused on diagnostics, the authors hardly report any accuracy measure and explicit description of the extraction process.

In the reviewed works, we found multiple disagreements on the numbers extracted from the primary studies. We found no data on many population features because of missing information and misleading or unclear statements, e.g. providing different information in text and figures and tables. Lack of scientific precision, e.g. reporting the *number of cases*, understood either as X-Ray images, or patients, without any discussion if the cases were symptomatic or not. Inconsistencies were also noticed in reporting of DL architecture. For instance, the following names were used across multiple studies: *ResNet-18, ResNet18, resnet-18, 18-layer ResNet*. In some of the papers, the architecture was not reported at all.

Some level of discrepancy was also observed in extracting the measurements of AI models performance. For example, the diagnostic effectiveness metrics from primary studies were either not reported or incorrectly reported by multiple studies. Such negligence may lead to further replicating the errors by subsequent studies and should be corrected before releasing the paper, or soon after in an updated version or erratum.

The amount of waste and poor biomedical research quality is staggering [70,82]. Papers that do not bring any additional knowledge to the field can be deemed redundant in line with multiple papers and International Committee of Medical Journal Editors (ICMJE) guidelines [83–86].

Many of the reviews included in this paper did not contribute to strengthening the evidence on using AI in diagnosing COVID-19 from medical imaging. Those works have not even identified and correctly cited preexisting studies, which is deemed the essence of any research. Some of the potential explanations are that multiple similar studies might have been initiated around the same time and prolonged review times impacted their content. Alternatively, the papers were too broadly focused and have not actually touched on application of AI to medical imaging. As the research questions might have not been strict, the reviews might have been focused on vast areas, e.g. using AI in the battle of COVID-19.

The low credibility of evidence, flawed reporting (e.g. on population characteristics), could be associated with lack of knowledge of reporting standards, clinical practice, or misunderstandings regarding AI techniques.

Proper reporting of deep learning performance from primary research studies is challenging. Naude [32] has pointed out some of the major concerns regarding adoption of AI in COVID-19 research, including data availability, sufficient quality of the data (lack of noise and outliers) as well as privacy protection. Still many studies do not share the source code as open source, which highly limits reproducibility of their findings [87, 88].

### 4.1 Study Strengths & Limitations

Our umbrella review has the following strengths. First, the search strategy was comprehensive based on adequate inclusion criteria related to research question, and spanned across a wide selection of existing data sources: papers and preprints. This selection was further expanded by searching the references of included papers to identify new works. Noteworthy, the searches were not limited in terms of format or language. The process of our review was rigorous as the study was proceeded by the publication of protocol. We used the most up-to-date and applicable tools to assess credibility and quality of reporting – AMSTAR 2 and PRISMA with extension for DTA, respectively. Nevertheless, these instruments have been designed for reviews in the fields of medicine and health sciences, where the formulation of the research question is structured, the methodology is validated and the quantitative synthesis of results is more popular. There are also other limitations associated with this study. Although our exhaustive and sensitive search covered multiple aspects, some studies might have still been missed. We did not search for Chinese data sources that could include many valuable papers. Secondly, it must be noted that the vast majority of the included studies focused on a broader context than purely diagnosing COVID-19 from medical images.

In this study, we have investigated wasting among the reviews. We based on the date of publishing of the last included primary study in particular review. This approach aims at assessing the depth of the search performed by the authors. We assumed that if the authors included a given study, they should have had the required knowledge about all the studies that appeared prior that date. This approach relaxes the strict requirement to include all the studies that appeared before the review was published and seems to more objectively measure the quality of the search. Due to the lack of validated method, we based this approach on intuition.

This study does not include quantitative analysis and extended credibility evaluation, as this is planned to be addressed in the follow-up papers. Thus, we also do not present a subgroup analysis and investigation of heterogeneity nor sensitivity analysis.

## 5 Conclusions

The COVID-19 research is quickly moving forward, each day hundreds of new papers are published. As AI starts to play increasingly important role in clinical practice [89, 90], it is crucial to properly evaluate its performance using well established standards.

In this paper, we synthesised and assessed the quality of the 22 reviews that mention using AI on COVID-19 medical images. We reviewed them and critically assessed their quality using well established instruments from PRISMA-DTA [44] and AMSTAR 2 [43].

The overall conclusion from this study is that we still need high quality evidence to prove the effectiveness of AI in detecting COVID-19 from medical images. The recommendation on using AI to detect COVID-19 in medical images can not be provided until more credible studies are presented, and the certainty of evidence becomes evaluated using well established techniques.

In order to accomplish this, we urge the authors of the systematic reviews to use PRISMA and AMSTAR 2, and the authors of primary studies to adapt the *Checklist for Artificial Intelligence in Medical Imaging (CLAIM)* – a new standard for transparency and reproducibility of the research [91].

It is high time to adapt best practices and improve the quality of the research, as well as to apply higher scrutiny in filtering out nonconstructive contributions.

## Data Availability

Supplementary material is appended to the end of the main paper.

## Authors contributions

We followed CRediT (Contributor Roles Taxonomy) to report on authors contributions.

1. Paweł Jemioło (PJ): Conceptualisation, Validation, Formal analysis, Investigation, Data Curation, Writing – Original Draft, Writing – Review & Editing, Visualisation, Project administration.
2. Dawid Storman (DS): Conceptualisation, Methodology, Validation, Formal analysis, Investigation, Writing – Original Draft, Writing – Review & Editing, Visualisation.
3. Patryk Orzechowski (PO): Conceptualisation, Investigation, Software, Visualisation, Writing – Original Draft, Writing – Review & Editing.

## Acknowledgement

The authors would like to thank Prof. Jason H. Moore from the University of Pennsylvania for his kind support and useful suggestions.

## Data and Code Availability

Full extraction forms and code used in this research can be accessed via contact with corresponding author.

## A Search Strategies

### A.1 MEDLINE

1. exp Coronavirus Infections/ or exp Coronavirus/ or exp Betacoronavirus/ or (“Coronavirus Infection*” or “Coronavirus” or “Betacoronavirus”).ti,ab. or ((“corona*” or “corono*”) adj1 (“virus*” or “viral*” or “virinae*”)).ti,ab. or (“coronavirus*” or “Severe acute respiratory syndrome related coronavirus” or “Severe acute respiratory syndrome coronavirus 2” or “coronovirus*” or “coron?virinae*” or “2019-nCoV” or “2019nCoV” or “2019-CoV” or “nCoV2019” or “nCoV-2019” or “COVID-19” or “COVID19” or “CORVID-19” or “CORVID19” or “WN-CoV” or “WNCoV” or “HCoV-19” or “HCoV19” or “CoV” or “2019 novel*” or “2019 novel coronavirus” or “2019 nCoV” or “Ncov” or “n-cov” or “SARS-CoV-2” or “SARSCoV-2” or “SARSCoV2” or “SARS-CoV2” or “SARSCov19” or “SARS-Cov19” or “SARSCov-19” or “SARS-Cov-19” or “SARSr-cov” or “Ncovor” or “Ncorona*” or “Ncorono*” or “NcovWuhan*” or “NcovHubei*” or “NcovChina*” or “NcovChinese*” or “Wuhan virus*” or “novel CoV” or “CoV 2” or “CoV2” or “betacoron?vir*”).ti,ab. or (((“respiratory*” adj2 (“acute*” or “symptom*” or “disease*” or “ill-ness*” or “condition*”)) or “sea-food market*” or “seafood market*” or “food market*” or “foodmarket*”) adj10 (“Wuhan*” or “Hubei*” or “China*” or “Chinese*” or “Huanan*”)).ti,ab. or ((“outbreak*” or “wildlife*” or “wild-life” or “pandemic*” or “epidemic*”) adj3 (“Wuhan*” or “Hubei*” or “China*” or “Chinese*” or “Huanan*”)).ti,ab. or (“anti-flu*” or “anti-influenza*” or “antiflu*” or “antinfluenza*”).ti,ab.
2. (“influenza” or “AIDS” or “immunodeficiency virus” or “HIV” or “sexually transmitted disease” or “sexually transmitted infections” or “STD” or “STI”).ti,ab.
3. (“recogni*” or “classif*” or “regress*” or “clusteri*” or “discriminat*” or “detect*” or “categori*” or “estimat*”).ti,ab.
4. (“Machine Learning” or “DL” or “Deep Learning” or “Representation Learning” or “Transfer Learning” or “AI” or “Artificial intelligen*” or “Computational Intelligen*”).ti,ab.
5. (“MLP” or ((“multi-layer” or “multi layer”) and “perceptron”) or “LSTM” or “BLSTM” or “GAN” or “generative adversarial” or “RNN” or “ANN” or “DNN” or “CNN” or “NN” or “Neural Network*” or “SVM” or “SVC” or “support vector*” or “LDA” or “QDA” or “discriminant analysis” or “naive bayes*” or “knn” or “nearest neighb*” or “Decision*” or “Expert*” or ((“Logistic” or “Linear”) AND “Regress*”) or “Random Forest” or “Gradient Boost*” or “AdaBoost” or “XGBoost” or “LightGBM” or “classifier*” or “regressor*”).ti,ab.
6. exp diagnostic imaging/ or exp diagnosis computer assisted/ or exp Tomography, Emission Computed/ or exp Tomography, X-ray computed/ or exp echography/ or exp magnetic resonance imaging/ or (“diagnostic imaging” or “computer assisted” or “computer-assisted” or “Tomography” or “Emission Computed” or “Emission-Computed” or “X-ray computed” or “X ray computed” or “X-ray-computed” or “echography” or “magnetic resonance imaging” or “mri” or “magnetic resonance imaging” or “microscop*” or “photograph*” or “holograph*” or “radiograph*” or “spectroscop*” or “stroboscop*” or “subtraction technique*” or “thermograph*” or “tomograph*” or “transilluminat*” or “ultrasono-graph*” or “ultrasound” or “imaging” or “scan*” or “X-Ray” or “X Ray” or “CT Scan” or “Computed Tomography” or “CT” or “PET” or “PET-CT” or “positron emission tomograph*” or “MRI” or “fMRI” or “NMRI” or “scintigraph*” or “Doppler echography” or “sonograph*” or “ultraso*” or “doppler” or “magnetic resonance imag*”).ti,ab.
7. (“overview*” or “review*” or “survey*”).af. or exp review/
8. (1 not 2) and (3 or 4 or 5) and 6 and 7

### A.2 EMBASE

1. ‘coronavirus infections’/exp OR ‘coronavirus’/exp OR ‘betacoronavirus’/exp OR ‘coronavirus infection*’:ti,ab OR ‘coronavirus’:ti,ab OR ‘betacoronavirus’:ti,ab OR (((‘corona*’ OR ‘corono*’) NEAR/1 (‘virus*’ OR ‘viral*’ OR ‘virinae*’)):ti,ab) OR ‘coronavirus*’:ti,ab OR ‘severe acute respiratory syndrome related coronavirus’:ti,ab OR ‘severe acute respiratory syndrome coronavirus 2’:ti,ab OR ‘coronovirus*’:ti,ab OR ‘coron?virinae*’:ti,ab OR ‘2019-ncov’:ti,ab OR ‘2019ncov’:ti,ab OR ‘2019-cov’:ti,ab OR ‘ncov2019’:ti,ab OR ‘ncov-2019’:ti,ab OR ‘covid-19’:ti,ab OR ‘covid19’:ti,ab OR ‘corvid-19’:ti,ab OR ‘corvid19’:ti,ab OR ‘wn-cov’:ti,ab OR ‘wncov’:ti,ab OR ‘hcov-19’:ti,ab OR ‘hcov19’:ti,ab OR ‘cov’:ti,ab OR ‘2019 novel*’:ti,ab OR ‘2019 novel coronavirus’:ti,ab OR ‘2019 ncov’:ti,ab OR ‘ncov’:ti,ab OR ‘n-cov’:ti,ab OR ‘sars-cov-2’:ti,ab OR ‘sarscov-2’:ti,ab OR ‘sarscov2’:ti,ab OR ‘sars-cov2’:ti,ab OR ‘sarscov19’:ti,ab OR ‘sars-cov19’:ti,ab OR ‘sarscov-19’:ti,ab OR ‘sars-cov-19’:ti,ab OR ‘sarsr-cov’:ti,ab OR ‘ncovor’:ti,ab OR ‘ncorona*’:ti,ab OR ‘ncorono*’:ti,ab OR ‘ncovwuhan*’:ti,ab OR ‘ncovhubei*’:ti,ab OR ‘ncovchina*’:ti,ab OR ‘ncovchinese*’:ti,ab OR ‘wuhan virus*’:ti,ab OR ‘novel cov’:ti,ab OR ‘cov 2’:ti,ab OR ‘cov2’:ti,ab OR ‘betacoron?vir*’:ti,ab OR (((‘outbreak*’ OR ‘wildlife*’ OR ‘wild-life’ OR ‘pandemic*’ OR ‘epidemic*’) NEAR/3 (‘wuhan*’ OR ‘hubei*’ OR ‘china*’ OR ‘chinese*’ OR ‘hua-nan*’)):ti,ab) OR ‘anti-flu*’:ti,ab OR ‘anti-influenza*’:ti,ab OR ‘antiflu*’:ti,ab OR ‘antinfluenza*’:ti,ab OR (((‘respiratory*’ OR ‘sea-food market*’ OR ‘seafood market*’ OR ‘food market*’ OR ‘foodmarket*’) NEAR/10 (‘wuhan*’ OR ‘hubei*’ OR ‘china*’ OR ‘chinese*’ OR ‘huanan*’)):ti,ab)
2. ‘influenza’:ti,ab OR ‘aids’:ti,ab OR ‘immunodeficiency virus’:ti,ab OR ‘hiv’:ti,ab OR ‘sexually transmitted disease’:ti,ab OR ‘sexually transmitted infections’:ti,ab OR ‘std’:ti,ab OR ‘sti’:ti,ab
3. ‘recogni*’:ti,ab OR ‘classif*’:ti,ab OR ‘regress*’:ti,ab OR ‘clusteri*’:ti,ab OR ‘discriminat*’:ti,ab OR ‘detect*’:ti,ab OR ‘categori*’:ti,ab OR ‘estimat*’:ti,ab
4. ‘machine learning’:ti,ab OR ‘dl’:ti,ab OR ‘deep learning’:ti,ab OR ‘representation learning’:ti,ab OR ‘transfer learning’:ti,ab OR ‘ai’:ti,ab OR ‘artificial intelligen*’:ti,ab OR ‘computational intelligen*’:ti,ab
5. ‘mlp’:ti,ab OR ((‘multi-layer’:ti,ab OR ‘multi layer’:ti,ab) AND ‘perceptron’:ti,ab) OR ‘lstm’:ti,ab OR ‘blstm’:ti,ab OR ‘gan’:ti,ab OR ‘generative adversarial’:ti,ab OR ‘rnn’:ti,ab OR ‘ann’:ti,ab OR ‘dnn’:ti,ab OR ‘cnn’:ti,ab OR ‘nn’:ti,ab OR ‘neural network*’:ti,ab OR ‘svm’:ti,ab OR ‘svc’:ti,ab OR ‘support vector*’:ti,ab OR ‘lda’:ti,ab OR ‘qda’:ti,ab OR ‘discriminant analysis’:ti,ab OR ‘naive bayes*’:ti,ab OR ‘knn’:ti,ab OR ‘nearest neighb*’:ti,ab OR ‘decision*’:ti,ab OR ‘expert*’:ti,ab OR ((‘logistic’:ti,ab OR ‘linear’:ti,ab) AND ‘regress*’:ti,ab) OR ‘random forest’:ti,ab OR ‘gradient boost*’:ti,ab OR ‘adaboost’:ti,ab OR ‘xgboost’:ti,ab OR ‘lightgbm’:ti,ab OR ‘classifier*’:ti,ab OR ‘regressor*’:ti,ab
6. ‘diagnostic imaging’/exp OR ‘diagnosis computer assisted’/exp OR ‘tomography, emission computed’ /exp OR ‘tomography, x-ray computed’/exp OR ‘echography’/exp OR ‘magnetic resonance imaging’/exp OR ‘diagnostic imaging’:ti,ab OR ‘computer assisted’:ti,ab OR ‘computer-assisted’:ti,ab OR ‘tomography’:ti,ab OR ‘emission computed’:ti,ab OR ‘emission-computed’:ti,ab OR ‘x-ray computed’: ti,ab OR ‘x ray computed’:ti,ab OR ‘x-ray-computed’:ti,ab OR ‘echography’:ti,ab OR ‘magnetic resonance imaging’:ti,ab OR ‘microscop*’:ti,ab OR ‘photograph*’:ti,ab OR ‘holograph*’:ti,ab OR ‘radiograph*’:ti,ab OR ‘spectroscop*’:ti,ab OR ‘stroboscop*’:ti,ab OR ‘subtraction technique*’:ti,ab OR ‘thermograph*’:ti,ab OR ‘tomograph*’:ti,ab OR ‘transilluminat*’:ti,ab OR ‘ultrasonograph*’:ti,ab OR ‘ultrasound’:ti,ab OR ‘imaging’:ti,ab OR ‘scan*’:ti,ab OR ‘x-ray’:ti,ab OR ‘x ray’:ti,ab OR ‘ct scan’:ti,ab OR ‘computed tomography’:ti,ab OR ‘ct’:ti,ab OR ‘pet’:ti,ab OR ‘pet-ct’:ti,ab OR (‘positron’:ti,ab AND ‘emission’:ti,ab AND ‘tomograph*’:ti,ab) OR ‘mri’:ti,ab OR ‘fmri’:ti,ab OR ‘nmri’:ti,ab OR ‘scintigraph*’:ti,ab OR (‘doppler’:ti,ab AND ‘echography’:ti,ab) OR ‘sonograph*’:ti,ab OR ‘ultraso*’: ti,ab OR ‘doppler’:ti,ab OR (‘magnetic’:ti,ab AND ‘resonance’:ti,ab AND ‘imag*’:ti,ab)
7. overview* OR review* OR survey* OR ‘review’/exp
8. (#1 not #2) and (#3 or #4 or #5) and #6 and #7

### A.3 Web of Science

1. TS=(‘Coronavirus Infection*’ or ‘Coronavirus’ or ‘Betacoronavirus’) or TS=((‘corona*’ or ‘corono*’) NEAR/1 (‘virus*’ or ‘viral*’ or ‘virinae*’)) or TS=(‘coronavirus*’ or ‘Severe acute respiratory syndrome related coronavirus’ or ‘Severe acute respiratory syndrome coronavirus 2’ or ‘coronovirus*’ or ‘coron?virinae*’ or ‘2019-nCoV’ or ‘2019nCoV’ or ‘2019-CoV’ or ‘nCoV2019’ or ‘nCoV-2019’ or ‘COVID-19’ or ‘COVID19’ or ‘CORVID-19’ or ‘CORVID19’ or ‘WN-CoV’ or ‘WNCoV’ or ‘HCoV-19’ or ‘HCoV19’ or ‘CoV’ or ‘2019 novel*’ or ‘2019 novel coronavirus’ or ‘2019 nCoV’ or ‘Ncov’ or ‘n-cov’ or ‘SARS-CoV-2’ or ‘SARSCoV-2’ or ‘SARSCoV2’ or ‘SARS-CoV2’ or ‘SARSCov19’ or ‘SARS-Cov19’ or ‘SARSCov-19’ or ‘SARS-Cov-19’ or ‘SARSr-cov’ or ‘Ncovor’ or ‘Ncorona*’ or ‘Ncorono*’ or ‘Ncov-Wuhan*’ or ‘NcovHubei*’ or ‘NcovChina*’ or ‘NcovChinese*’ or ‘Wuhan virus*’ or ‘novel CoV’ or ‘CoV 2’ or ‘CoV2’ or ‘betacoron?vir*’) or TS=(((‘respiratory*’ NEAR/2 (‘acute*’ or ‘symptom*’ or ‘disease*’ or ‘illness*’ or ‘condition*’)) or ‘sea-food market*’ or ‘seafood market*’ or ‘food market*’ or ‘foodmarket*’) NEAR/10 (‘Wuhan*’ or ‘Hubei*’ or ‘China*’ or ‘Chinese*’ or ‘Huanan*’)) or TS=((‘outbreak*’ or ‘wildlife*’ or ‘wild-life’ or ‘pandemic*’ or ‘epidemic*’) NEAR/3 (‘Wuhan*’ or ‘Hubei*’ or ‘China*’ or ‘Chinese*’ or ‘Huanan*’)) or TS=(‘anti-flu*’ or ‘anti-influenza*’ or ‘antiflu*’ or ‘antinfluenza*’)
2. TS=(‘influenza’ or ‘AIDS’ or ‘immunodeficiency virus’ or ‘HIV’ or ‘sexually transmitted disease’ or ‘sexually transmitted infections’ or ‘STD’ or ‘STI’)
3. TS=(‘recogni*’ or ‘classif*’ or ‘regress*’ or ‘clusteri*’ or ‘discriminat*’ or ‘detect*’ or ‘categori*’ or ‘estimat*’)
4. TS=(‘Machine Learning’ or ‘DL’ or ‘Deep Learning’ or ‘Representation Learning’ or ‘Transfer Learning’ or ‘AI’ or ‘Artificial intelligen*’ or ‘Computational Intelligen*’)
5. TS=(‘MLP’ or ((‘multi-layer’ or ‘multi layer’) and ‘perceptron’) or ‘LSTM’ or ‘BLSTM’ or ‘GAN’ or ‘generative adversarial’ or ‘RNN’ or ‘ANN’ or ‘DNN’ or ‘CNN’ or ‘NN’ or ‘Neural Network*’ or ‘SVM’ or ‘SVC’ or ‘support vector*’ or ‘LDA’ or ‘QDA’ or ‘discriminant analysis’ or ‘naive bayes*’ or ‘knn’ or ‘nearest neighb*’ or ‘Decision*’ or ‘Expert*’ or ((‘Logistic’ or ‘Linear’) AND ‘Regress*’) or ‘Random Forest’ or ‘Gradient Boost*’ or ‘AdaBoost’ or ‘XGBoost’ or ‘LightGBM’ or ‘classifier*’ or ‘regressor*’)
6. TS=(‘diagnostic imaging’ or ‘computer assisted’ or ‘computer-assisted’ or ‘Tomography’ or ‘Emission Computed’ or ‘Emission-Computed’ or ‘X-ray computed’ or ‘X ray computed’ or ‘X-ray-computed’ or ‘echography’ or ‘magnetic resonance imaging’ or ‘mri’ or ‘magnetic resonance imaging’ or ‘microscop*’ or ‘photograph*’ or ‘holograph*’ or ‘radiograph*’ or ‘spectroscop*’ or ‘stroboscop*’ or ‘subtraction technique*’ or ‘thermograph*’ or ‘tomograph*’ or ‘transilluminat*’ or ‘ultrasonograph*’ or ‘ultrasound’ or ‘imaging’ or ‘scan*’ or ‘X-Ray’ or ‘X Ray’ or ‘CT Scan’ or ‘Computed Tomography’ or ‘CT’ or ‘PET’ or ‘PET-CT’ or ‘positron emission tomograph*’ or ‘MRI’ or ‘fMRI’ or ‘NMRI’ or ‘scintigraph*’ or ‘Doppler echography’ or ‘sonograph*’ or ‘ultraso*’ or ‘doppler’ or ‘magnetic resonance imag*’)
7. ALL=(‘overview*’ or ‘review*’ or ‘survey*’)
8. (#1 not #2) and (#3 or #4 or #5) and #6 and #7

### A.4 Scopus

((TITLE-ABS-KEY (“Coronavirus Infection*” OR “Coronavirus” OR “Betacoronavirus”) OR TITLE-ABSKEY ((“corona*” OR “corono*”) W/1 (“virus*” OR “viral*” OR “virinae*”)) OR TITLE-ABS-KEY (“coronavirus*” OR “Severe acute respiratory syndrome related coronavirus” OR “Severe acute respiratory syndrome coronavirus 2” OR “coronovirus*” OR “coron?virinae*” OR “2019-nCoV” OR “2019nCoV” OR “2019-CoV” OR “nCoV2019” OR “nCoV-2019” OR “COVID-19” OR “COVID19” OR “CORVID-19” OR “CORVID19” OR “WN-CoV” OR “WNCoV” OR “HCoV-19” OR “HCoV19” OR “CoV” OR “2019 novel*” OR “2019 novel coronavirus” OR “2019 nCoV” OR “Ncov” OR “n-cov” OR “SARS-CoV-2” OR “SARSCoV-2” OR “SARSCoV2” OR “SARS-CoV2” OR “SARSCov19” OR “SARS-Cov19” OR “SARSCov-19” OR “SARS-Cov-19” OR “SARSr-cov” OR “Ncovor” OR “Ncorona*” OR “Ncorono*” OR “NcovWuhan*” OR “NcovHubei*” OR “NcovChina*” OR “NcovChinese*” OR “Wuhan virus*” OR “novel CoV” OR “CoV 2” OR “CoV2” OR “betacoron?vir*”) OR TITLE-ABS-KEY (((“respiratory*” W/2 (“acute*” OR “symptom*” OR “disease*” OR “illness*” OR “condition*”)) OR “sea-food market*” OR “seafood market*” OR “food market*” OR “foodmarket*”) W/10 (“Wuhan*” OR “Hubei*” OR “China*” OR “Chinese*” OR “Huanan*”)) OR TITLE-ABS-KEY ((“outbreak*” OR “wildlife*” OR “wild-life” OR “pandemic*” OR “epidemic*”) W/3 (“Wuhan*” OR “Hubei*” OR “China*” OR “Chinese*” OR “Huanan*”)) OR TITLEABS-KEY (“anti-flu*” OR “anti-influenza*” OR “antiflu*” OR “antinfluenza*”)) AND NOT (TITLE-ABSKEY (“influenza” OR “AIDS” OR “immunodeficiency virus” OR “HIV” OR “sexually transmitted disease” OR “sexually transmitted infections” OR “STD” OR “STI”))) AND ((TITLE-ABS-KEY (“recogni*” OR “classif*” OR “regress*” OR “clusteri*” OR “discriminat*” OR “detect*” OR “categori*” OR “estimat*”)) OR (TITLE-ABS-KEY (“Machine Learning” OR “DL” OR “Deep Learning” OR “Representation Learning” OR “Transfer Learning” OR “AI” OR “Artificial intelligen*” OR “Computational Intelligen*”)) OR (TITLE-ABS-KEY (“MLP” OR “multi-layer perceptron” OR “multi layer perceptron” OR “LSTM” OR “BLSTM” OR “GAN” OR “generative adversarial” OR “RNN” OR “ANN” OR “DNN” OR “CNN” OR “NN” OR “Neural Network*” OR “SVM” OR “SVC” OR “support vector*” OR “LDA” OR “QDA” OR “discriminant analysis” OR “naive bayes*” OR “knn” OR “nearest neighb*” OR “Decision*” OR “Expert*” OR “Logistic Regress*” OR “Linear Regress*” OR “Random Forest” OR “Gradient Boost*” OR “AdaBoost” OR “XGBoost” OR “LightGBM” OR “classifier*” OR “regressor*”))) AND (TITLE-ABS-KEY (“diagnostic imaging” OR “computer assisted” OR “computer-assisted” OR “Tomography” OR “Emission Computed” OR “Emission-Computed” OR “X-ray computed” OR “X ray computed” OR “X-ray-computed” OR “echography” OR “magnetic resonance imaging” OR “mri” OR “magnetic resonance imaging” OR “microscop*” OR “photograph*” OR “holograph*” OR “radiograph*” OR “spectroscop*” OR “stroboscop*” OR “subtraction technique*” OR “thermograph*” OR “tomograph*” OR “transilluminat*” OR “ultrasonograph*” OR “ultrasound” OR “imaging” OR “scan*” OR “X-Ray” OR “X Ray” OR “CT Scan” OR “Computed Tomography” OR “CT” OR “PET” OR “PET-CT” OR “positron emission tomograph*” OR “MRI” OR “fMRI” OR “NMRI” OR “scintigraph*” OR “Doppler echography” OR “sonograph*” OR “ultraso*” OR “doppler” OR “magnetic resonance imag*”)) AND (ALL(“overview*” OR “review*” OR “survey*”))

### A.5 Cochrane Library

1. MeSH descriptor: [Coronavirus Infections] explode all trees
2. MeSH descriptor: [Coronavirus] explode all trees
3. MeSH descriptor: [Betacoronavirus] explode all trees
4. “coronavirus*” or “Severe acute respiratory syndrome related coronavirus” or “Severe acute respiratory syndrome coronavirus 2” or “coronovirus*” or “coron?virinae*” or “2019-nCoV” or “2019nCoV” or “2019-CoV” or “nCoV2019” or “nCoV-2019” or “COVID-19” or “COVID19” or “CORVID-19” or “CORVID19” or “WN-CoV” or “WNCoV” or “HCoV-19” or “HCoV19” or “CoV” or “2019 novel*” or “2019 novel coronavirus” or “2019 nCoV” or “Ncov” or “n-cov” or “SARS-CoV-2” or “SARSCoV-2” or “SARSCoV2” or “SARS-CoV2” or “SARSCov19” or “SARS-Cov19” or “SARSCov-19” or “SARS-Cov-19” or “SARSr-cov” or “Ncovor” or “Ncorona*” or “Ncorono*” or “NcovWuhan*” or “NcovHubei*” or “NcovChina*” or “NcovChinese*” or “Wuhan virus*” or “novel CoV” or “CoV 2” or “CoV2” or “betacoron?vir*”
5. (((“respiratory*” NEAR/2 (“acute*” or “symptom*” or “disease*” or “illness*” or “condition*”)) or “sea-food market*” or “seafood market*” or “food market*” or “foodmarket*”) NEAR/10 (“Wuhan*” or “Hubei*” or “China*” or “Chinese*” or “Huanan*”))
6. ((“outbreak*” or “wildlife*” or “wild-life” or “pandemic*” or “epidemic*”) NEAR/3 (“Wuhan*” or “Hubei*” or “China*” or “Chinese*” or “Huanan*”))
7. (“anti-flu*” or “anti-influenza*” or “antiflu*” or “antinfluenza*”)
8. #1 OR #2 OR #3 OR #4 OR #5 OR #6 OR #7
9. “influenza” or “AIDS” or “immunodeficiency virus” or “HIV” or “sexually transmitted disease” or “sexually transmitted infections” or “STD” or “STI”
10. “recogni*” or “classif*” or “regress*” or “clusteri*” or “discriminat*” or “detect*” or “categori*” or “estimat*”
11. “Machine Learning” or “DL” or “Deep Learning” or “Representation Learning” or “Transfer Learning” or “AI” or “Artificial intelligen*” or “Computational Intelligen*”
12. “MLP” or ((“multi-layer” or “multi layer”) and “perceptron”) or “LSTM” or “BLSTM” or “GAN” or “generative adversarial” or “RNN” or “ANN” or “DNN” or “CNN” or “NN” or “Neural Network*” or “SVM” or “SVC” or “support vector*” or “LDA” or “QDA” or “discriminant analysis” or “naive bayes*” or “knn” or “nearest neighb*” or “Decision*” or “Expert*” or ((“Logistic” or “Linear”) AND “Regress*”) or “Random Forest” or “Gradient Boost*” or “AdaBoost” or “XGBoost” or “LightGBM” or “classifier*” or “regressor*”
13. MeSH descriptor: [Diagnostic Imaging] explode all trees
14. MeSH descriptor: [Diagnosis, Computer-Assisted] explode all trees
15. MeSH descriptor: [Tomography, X-Ray Computed] explode all trees
16. MeSH descriptor: [Tomography, Emission-Computed] explode all trees
17. MeSH descriptor: [Ultrasonography] explode all trees
18. MeSH descriptor: [Magnetic Resonance Imaging] explode all trees
19. #13 or #14 or #15 or #16 or #17 or #18
20. (“diagnostic imaging” or “computer assisted” or “computer-assisted” or “Tomography” or “Emission Computed” or “Emission-Computed” or “X-ray computed” or “X ray computed” or “X-ray-computed” or “echography” or “magnetic resonance imaging” or “mri” or “magnetic resonance imaging” or “microscop*” or “photograph*” or “holograph*” or “radiograph*” or “spectroscop*” or “stroboscop*” or “subtraction technique*” or “thermograph*” or “tomograph*” or “transilluminat*” or “ultrasono-graph*” or “ultrasound” or “imaging” or “scan*” or “X-Ray” or “X Ray” or “CT Scan” or “Computed Tomography” or “CT” or “PET” or “PET-CT” or “positron emission tomograph*” or “MRI” or “fMRI” or “NMRI” or “scintigraph*” or “Doppler echography” or “sonograph*” or “ultraso*” or “doppler” or “magnetic resonance imag*”)
21. ((“overview*” or “review*” or “survey*”))
22. MeSH descriptor: [Review] explode all trees
23. #21 or #22
24. (#8 not #9) and (#10 or #11 or #12) and (#19 or #20) and #23

### A.6 IEEE Xplore

((“Coronavirus Infection” OR “Coronavirus” OR “Betacoronavirus”) OR ((“corona” OR “corono”) NEAR/1 (“virus” OR “viral” OR “virinae”)) OR (“Abstract”:”coronavirus” OR “Severe acute respiratory syndrome related coronavirus” OR “Severe acute respiratory syndrome coronavirus 2” OR “coronovirus” OR “coron?virinae” OR “2019-nCoV” OR “2019nCoV” OR “2019-CoV” OR “nCoV2019” OR “nCoV-2019” OR “COVID-19” OR “COVID19” OR “CORVID-19” OR “CORVID19” OR “WN-CoV” OR “WNCoV” OR “HCoV-19” OR “HCoV19” OR “CoV” OR “2019 novel” OR “2019 novel coronavirus” OR “2019 nCoV” OR “Ncov” OR “n-cov” OR “SARS-CoV-2” OR “SARSCoV-2” OR “SARSCoV2” OR “SARS-CoV2” OR “SARSCov19” OR “SARS-Cov19” OR “SARSCov-19” OR “SARS-Cov-19” OR “SARSr-cov” OR “Ncovor” OR “Ncorona” OR “Ncorono” OR “NcovWuhan” OR “NcovHubei” OR “NcovChina” OR “NcovChinese” OR “Wuhan virus” OR “novel CoV” OR “CoV 2” OR “CoV2” OR “betacoron?vir”) OR (“Document title”:”coronavirus” OR “Severe acute respiratory syndrome related coronavirus” OR “Severe acute respiratory syndrome coronavirus 2” OR “coronovirus” OR “coron?virinae” OR “2019-nCoV” OR “2019nCoV” OR “2019-CoV” OR “nCoV2019” OR “nCoV-2019” OR “COVID-19” OR “COVID19” OR “CORVID-19” OR “CORVID19” OR “WN-CoV” OR “WNCoV” OR “HCoV-19” OR “HCoV19” OR “CoV” OR “2019 novel” OR “2019 novel coronavirus” OR “2019 nCoV” OR “Ncov” OR “n-cov” OR “SARS-CoV-2” OR “SARSCoV-2” OR “SARSCoV2” OR “SARS-CoV2” OR “SARSCov19” OR “SARS-Cov19” OR “SARSCov-19” OR “SARS-Cov-19” OR “SARSr-cov” OR “Ncovor” OR “Ncorona” OR “Ncorono” OR “NcovWuhan” OR “NcovHubei” OR “NcovChina” OR “NcovChinese” OR “Wuhan virus” OR “novel CoV” OR “CoV 2” OR “CoV2” OR “betacoron?vir”) OR (((“respiratory” NEAR/2 (“acute” OR “symptom” OR “disease” OR “illness” OR “condition”)) OR “sea-food market” OR “seafood market” OR “food market” OR “foodmarket”) NEAR/10 (“Wuhan” OR “Hubei” OR “China” OR “Chinese” OR “Huanan”)) OR ((“outbreak” OR “wildlife” OR “wild-life” OR “pandemic” OR “epidemic”) NEAR/3 (“Wuhan” OR “Hubei” OR “China” OR “Chinese” OR “Huanan”)) OR (“anti-flu” OR “anti-influenza” OR “antiflu” OR “antinfluenza”)) AND ((“overview*” OR “review*” OR “survey*”) OR “Mesh Terms”:”review”)

### A.7 dblp

1. review|overview|survey
2. covid|corona|corono|ncov|wuhan|sars|betaco|corvid|hcov|ncov|hubei|virus|antin|pandem

### A.8 arXiv

1. AND abstract=covid* OR coronavirus; AND abstract=review* OR overview* OR survey*
2. AND title=covid* OR coronavirus; AND abstract=review* OR overview* OR survey*
3. AND abstract=covid* OR coronavirus; AND title=review* OR overview* OR survey*
4. AND title=covid* OR coronavirus; AND title=review* OR overview* OR survey*

### A.9 OSF Preprints

((“Coronavirus Infection*” OR “Coronavirus” OR “Betacoronavirus” OR”corona*” OR “corono*” OR “coronavirus*” OR “Severe acute respiratory syndrome related coronavirus” OR “Severe acute respiratory syndrome coronavirus 2” OR “coronovirus*” OR “coron?virinae*” OR “2019-nCoV” OR “2019nCoV” OR “2019-CoV” OR “nCoV2019” OR “nCoV-2019” OR “COVID-19” OR “COVID19” OR “CORVID-19” OR “CORVID19” OR “WN-CoV” OR “WNCoV” OR “HCoV-19” OR “HCoV19” OR “CoV” OR “2019 novel*” OR “2019 novel coronavirus” OR “2019 nCoV” OR “Ncov” OR “n-cov” OR “SARS-CoV-2” OR “SARSCoV-2” OR “SARSCoV2” OR “SARS-CoV2” OR “SARSCov19” OR “SARS-Cov19” OR “SARSCov-19” OR “SARS-Cov-19” OR “SARSr-cov” OR “NcovOR “ OR “Ncorona*” OR “Ncorono*” OR “NcovWuhan*” OR “NcovHubei*” OR “NcovChina*” OR “NcovChinese*” OR “Wuhan virus*” OR “novel CoV” OR “CoV 2” OR “CoV2” OR “betacoron?vir*” OR “respiratory*” OR “sea-food market*” OR “seafood market*” OR “food market*” OR “foodmarket*” OR “outbreak*” OR “wildlife*” OR “wild-life” OR “pandemic*” OR “epidemic*” OR “anti-flu*” OR “anti-influenza*” OR “antiflu*” OR “antinfluenza*”) NOT (“influenza” OR “AIDS” OR “immunodeficiency virus” OR “HIV” OR “sexually transmitted disease” OR “sexually transmitted infections” OR “STD” OR “STI”)) AND (“review*” OR “overview*” OR “survey*”) AND (“diagnostic imaging” OR “computer assisted” OR “computer-assisted” OR “Tomography” OR “Emission Computed” OR “Emission-Computed” OR “X-ray computed” OR “X ray computed” OR “X-ray-computed” OR “echography” OR “magnetic resonance imaging” OR “mri” OR “magnetic resonance imaging” OR “microscop*” OR “photograph*” OR “holograph*” OR “radiograph*” OR “spectroscop*” OR “stroboscop*” OR “subtraction technique*” OR “thermograph*” OR “tomograph*” OR “transilluminat*” OR “ultrasonograph*” OR “ultrasound” OR “imaging” OR “scan*” OR “X-Ray” OR “X Ray” OR “CT Scan” OR “Computed Tomography” OR “CT” OR “PET” OR “PET-CT” OR “positron emission tomograph*” OR “MRI” OR “fMRI” OR “NMRI” OR “scintigraph*” OR “Doppler echography” OR “sonograph*” OR “ultraso*” OR “doppler” OR “magnetic resonance imag*”) AND (“recogni*” OR “classif*” OR “regress*” OR “clusteri*” OR “discriminat*” OR “detect*” OR “categori*” OR “estimat*” OR “Machine Learning” OR “DL” OR “Deep Learning” OR “Representation Learning” OR “Transfer Learning” OR “AI” OR “Artificial intelligen*” OR “Computational Intelligen*” OR “MLP” OR “multi-layer perceptron” OR “multi layer perceptron” OR “LSTM” OR “BLSTM” OR “GAN” OR “generative adversarial” OR “RNN” OR “ANN” OR “DNN” OR “CNN” OR “NN” OR “Neural Network*” OR “SVM” OR “SVC” OR “support vector*” OR “LDA” OR “QDA” OR “discriminant analysis” OR “naive bayes*” OR “knn” OR “nearest neighb*” OR “Decision*” OR “Expert*” OR “Logistic Regress*” OR “Linear Regress*” OR “Random Forest” OR “Gradient Boost*” OR “AdaBoost” OR “XGBoost” OR “LightGBM” OR “classifier*” OR “regressor*”)

## B Included studies

**Table 3:**
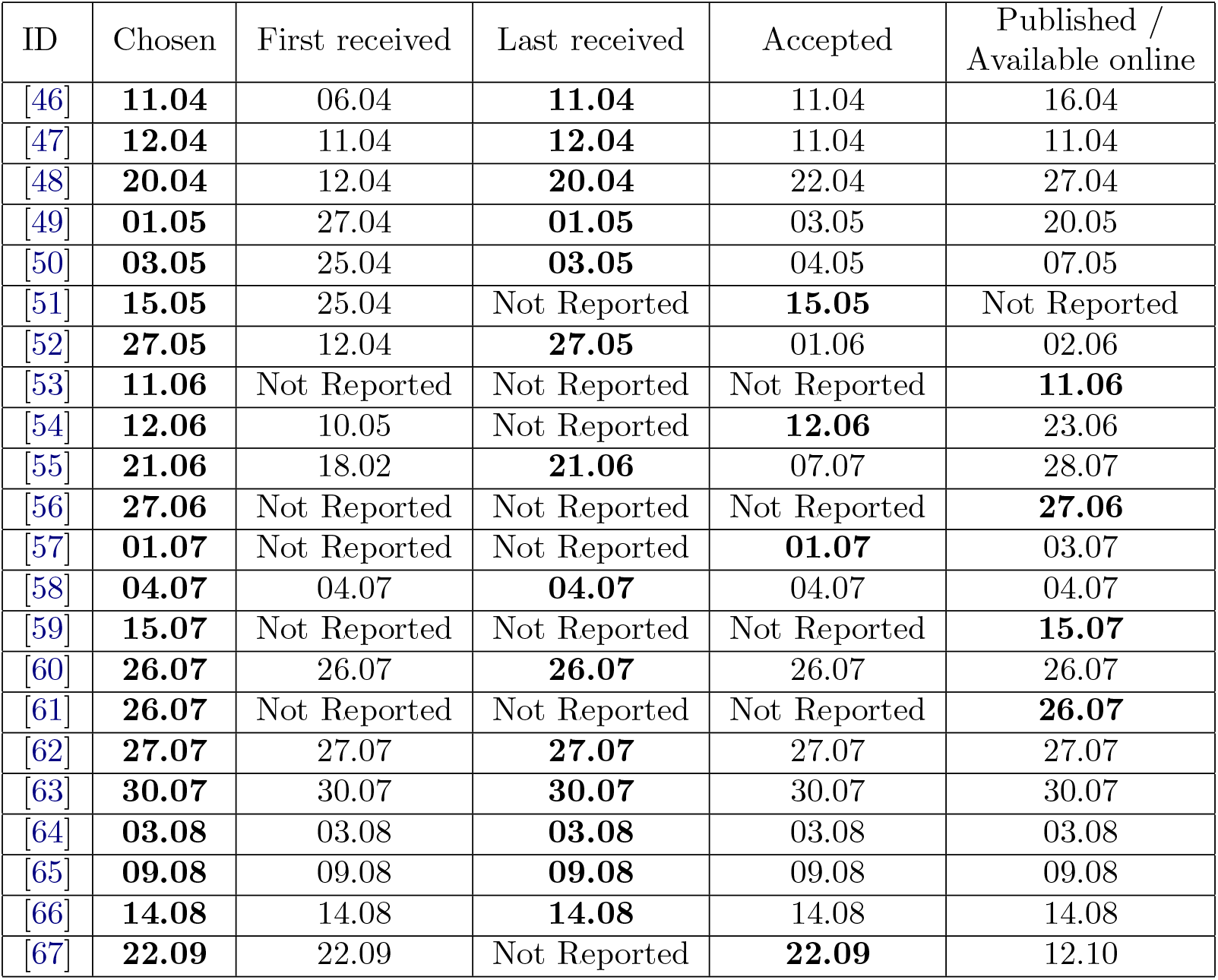
Included papers with dates.

| ID | Chosen | First received | Last received | Accepted | Published / Available online |
| --- | --- | --- | --- | --- | --- |
| [46] | **11.04** | 06.04 | **11.04** | 11.04 | 16.04 |
| [47] | **12.04** | 11.04 | **12.04** | 11.04 | 11.04 |
| [48] | **20.04** | 12.04 | **20.04** | 22.04 | 27.04 |
| [49] | **01.05** | 27.04 | **01.05** | 03.05 | 20.05 |
| [50] | **03.05** | 25.04 | **03.05** | 04.05 | 07.05 |
| [51] | **15.05** | 25.04 | Not Reported | **15.05** | Not Reported |
| [52] | **27.05** | 12.04 | **27.05** | 01.06 | 02.06 |
| [53] | **11.06** | Not Reported | Not Reported | Not Reported | **11.06** |
| [54] | **12.06** | 10.05 | Not Reported | **12.06** | 23.06 |
| [55] | **21.06** | 18.02 | **21.06** | 07.07 | 28.07 |
| [56] | **27.06** | Not Reported | Not Reported | Not Reported | **27.06** |
| [57] | **01.07** | Not Reported | Not Reported | **01.07** | 03.07 |
| [58] | **04.07** | 04.07 | **04.07** | 04.07 | 04.07 |
| [59] | **15.07** | Not Reported | Not Reported | Not Reported | **15.07** |
| [60] | **26.07** | 26.07 | **26.07** | 26.07 | 26.07 |
| [61] | **26.07** | Not Reported | Not Reported | Not Reported | **26.07** |
| [62] | **27.07** | 27.07 | **27.07** | 27.07 | 27.07 |
| [63] | **30.07** | 30.07 | **30.07** | 30.07 | 30.07 |
| [64] | **03.08** | 03.08 | **03.08** | 03.08 | 03.08 |
| [65] | **09.08** | 09.08 | **09.08** | 09.08 | 09.08 |
| [66] | **14.08** | 14.08 | **14.08** | 14.08 | 14.08 |
| [67] | **22.09** | 22.09 | Not Reported | **22.09** | 12.10 |

## C Excluded studies with reasons

**Table 4:** Exluded studies with reasons.

| ID | Reason |
| --- | --- |
| [92] | Wrong Study Design |
| [93] | Wrong Patient Population |
| [94] | Wrong Target Condition |
| [95] | Wrong Outcomes |
| [96] | Wrong Outcomes |
| [97] | Wrong Outcomes |
| [98] | Wrong Outcomes |
| [99] | Wrong Outcomes |
| [100] | Wrong Outcomes |
| [101] | Wrong Outcomes |
| [102] | Wrong Outcomes |

## D Full characteristics of included reviews

**Table 5:**
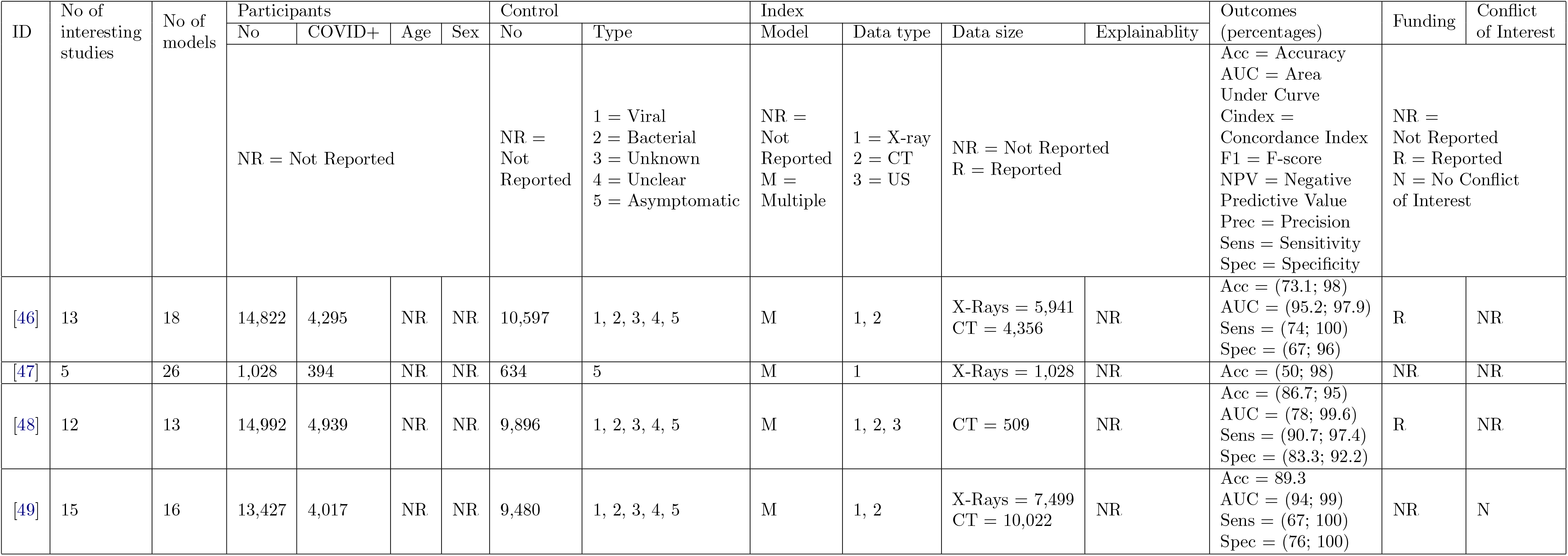
Full characteristics of included reviews, part 1.

**Table 6:**
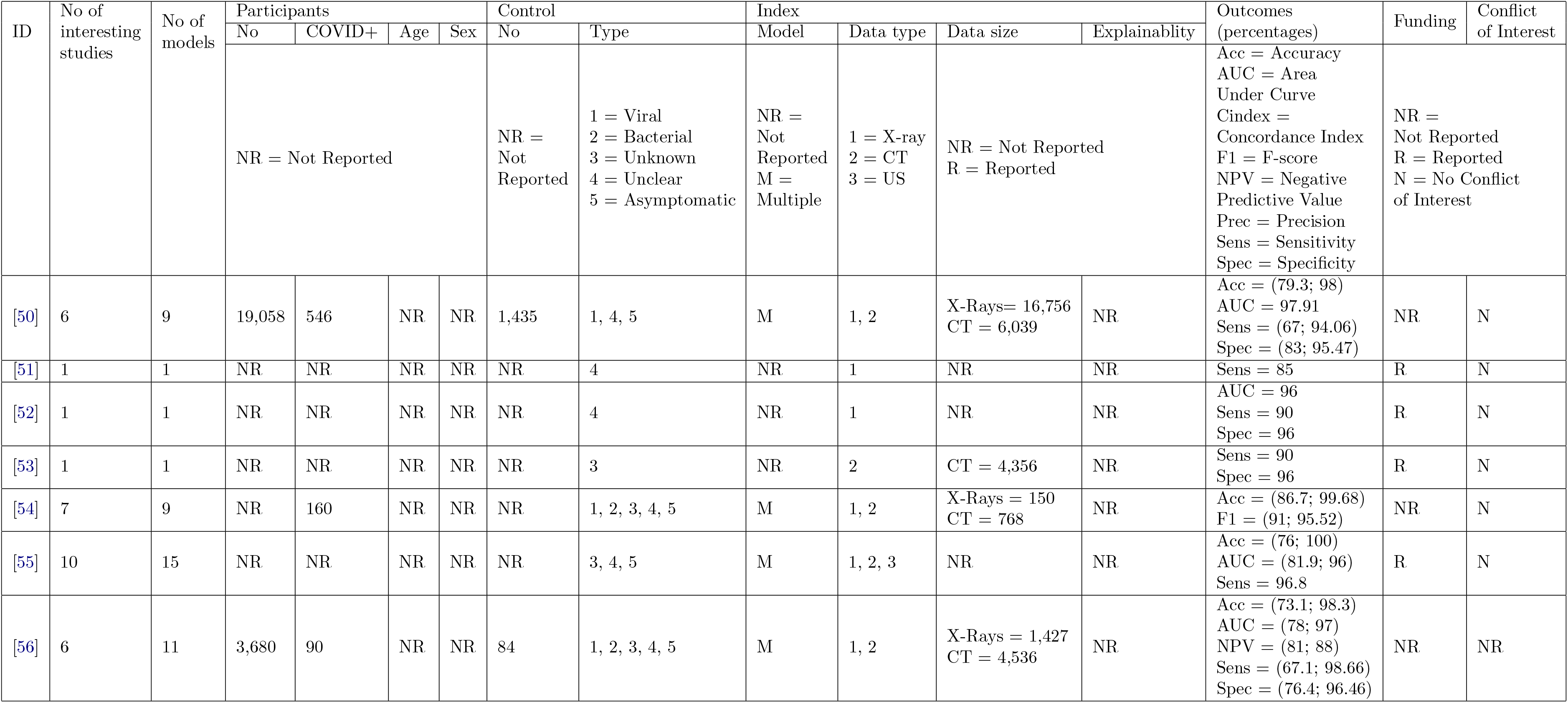
Full characteristics of included reviews, part 2.

**Table 7:**
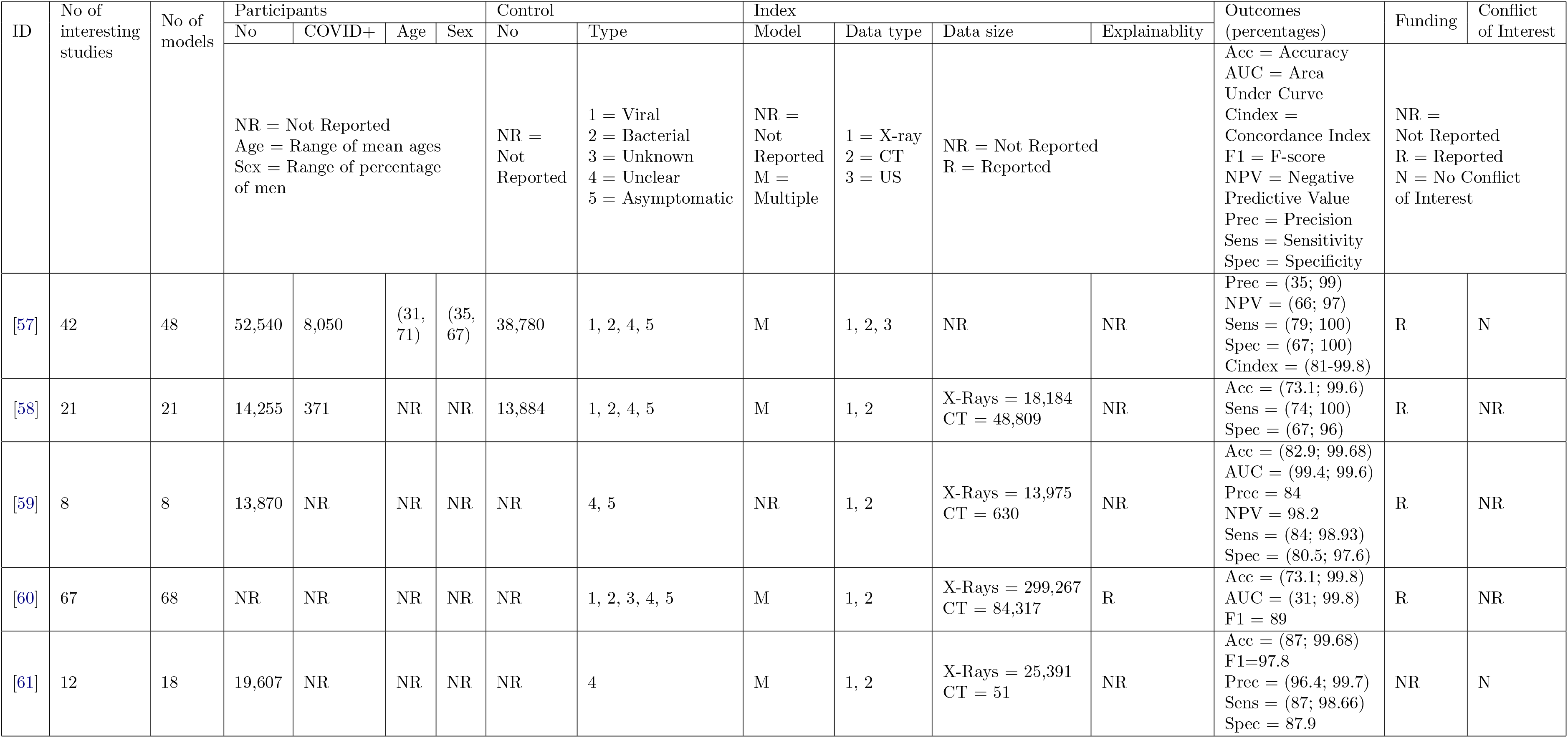
Full characteristics of included reviews, part 3.

**Table 8:**
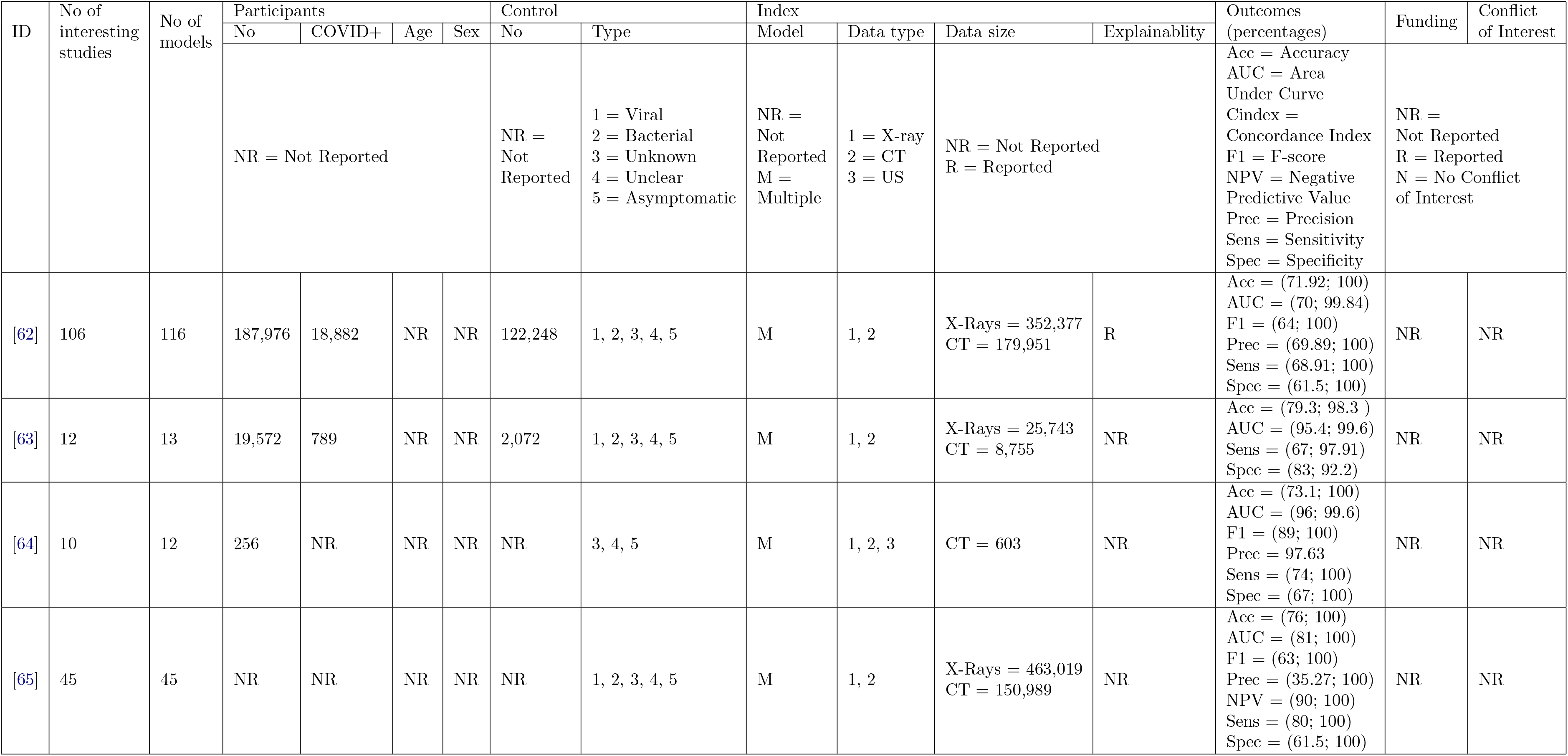
Full characteristics of included reviews, part 4.

**Table 9:** Full characteristics of included reviews, part 5.

| ID | No of interesting studies | No of models | Participants |  |  |  | Control |  | Index |  |  |  | Outcomes (percentages) | Funding | Conflict of Interest |
| --- | --- | --- | --- | --- | --- | --- | --- | --- | --- | --- | --- | --- | --- | --- | --- |
|  |  |  | No | COVID+ | Age | Sex | No | Type | Model | Data type | Data size | Explainability |  |  |  |
|  |  |  | NR = Not Reported |  |  |  | NR = Not Reported | 1 = Viral<br>2 = Bacterial<br>3 = Unknown<br>4 = Unclear<br>5 = Asymptomatic | NR = Not Reported<br>Not Reported<br>M = Multiple | 1 = X-ray<br>2 = CT<br>3 = US | NR = Not Reported<br>R = Reported |  | Acc = Accuracy<br>AUC = Area Under Curve<br>Cindex = Concordance Index<br>F1 = F-score<br>NPV = Negative Predictive Value<br>Prec = Precision<br>Sens = Sensitivity<br>Spec = Specificity | NR = Not Reported<br>Not Reported<br>R = Reported<br>N = No Conflict of Interest |  |
| [66] | 24 | 29 | 3,238 | 1,650 | NR | NR | 1,578 | 1, 2, 3, 4, 5 | M | 1, 2 | X-Rays = 32,840<br>CT = 149,078 | NR | Acc = (64.4; 99.87)<br>AUC = (67; 100)<br>F1 = (70; 99.8)<br>Prec = (89.8; 100)<br>NPV=100<br>Sens = (61; 100)<br>Spec = (28; 100) | R | N |
| [67] | 27 | 31 | 21,053 | 2,570 | NR | NR | 1,678 | 1, 2, 3, 4, 5 | M | 1, 2 | X-Rays = 194,470<br>CT = 55,245 | NR | Acc = (73.1; 99.68)<br>AUC (92; 99.6)<br>F1 = (87; 97.5)<br>NPV = (84; 93.36)<br>Sens = (8; 100)<br>Spec = (67; 99.99) | R | NR |

## E Primary studies per review

Table description: **A** – number of primary studies included in the review (see Figure 4); **D** – number of unique included primary papers (see Figure 4); **E** – cumulative sum of unique included primary papers; **J** – cited reviews that were published; **K** – cited reviews both published and available as preprints; **X** – all primary papers available to the date of publishing last included study; **Y** – all primary papers available to the chosen date (see Appendix B, see Figure 4)).

**Table 10:**
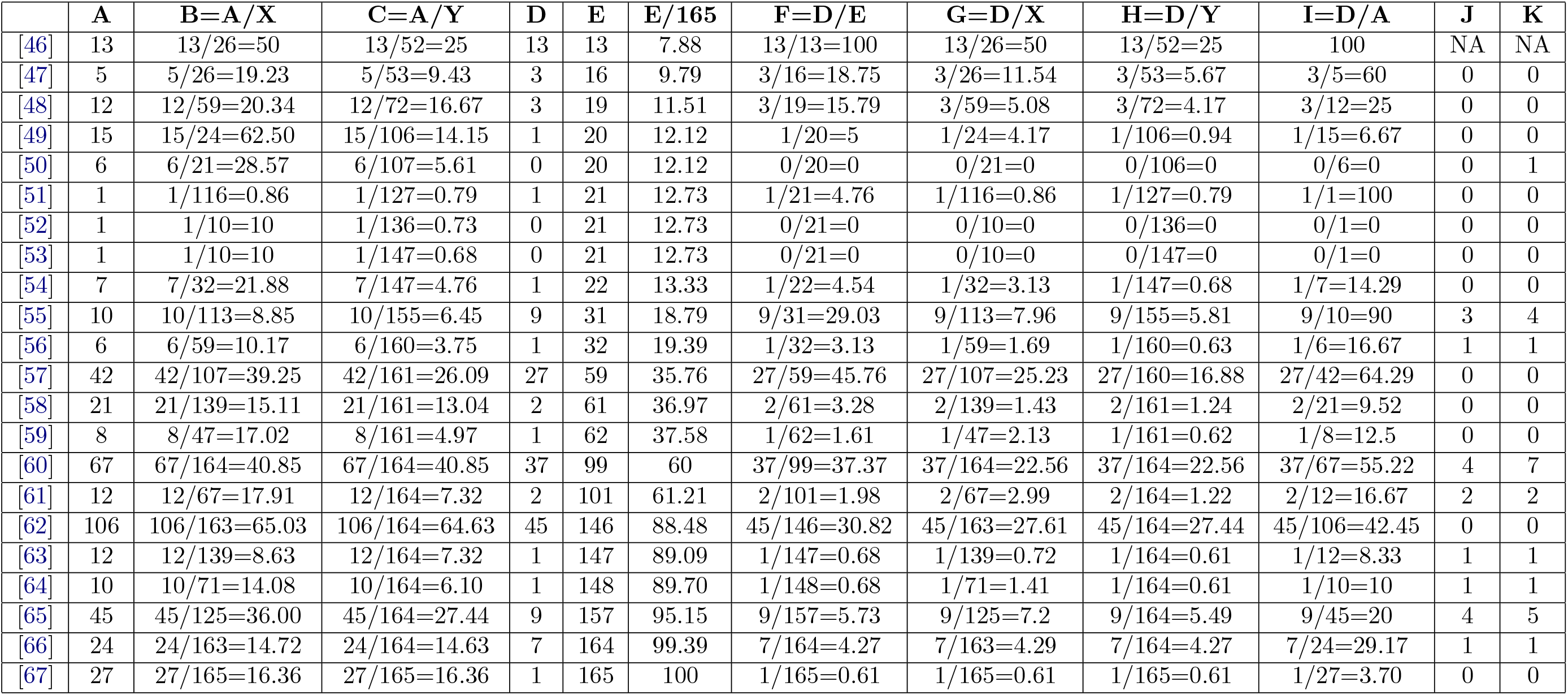
Time and resource wasting statistics.

## F PRISMA-DTA heatmap

**Figure 5:**
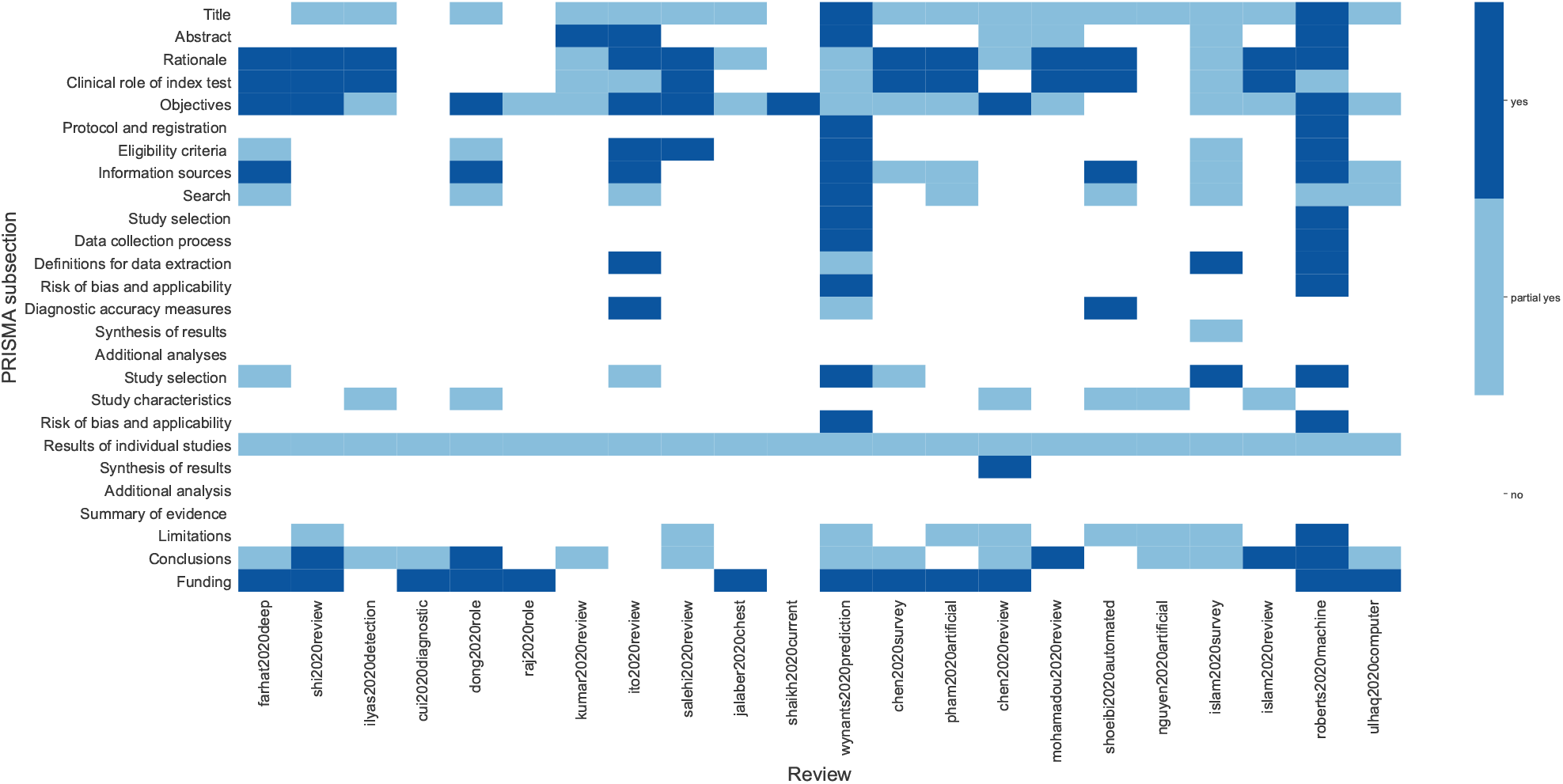
Review authors’ judgements about each PRISMA-DTA item across all included studies.

## G PRISMA-DTA results per review

**Figure 6:**
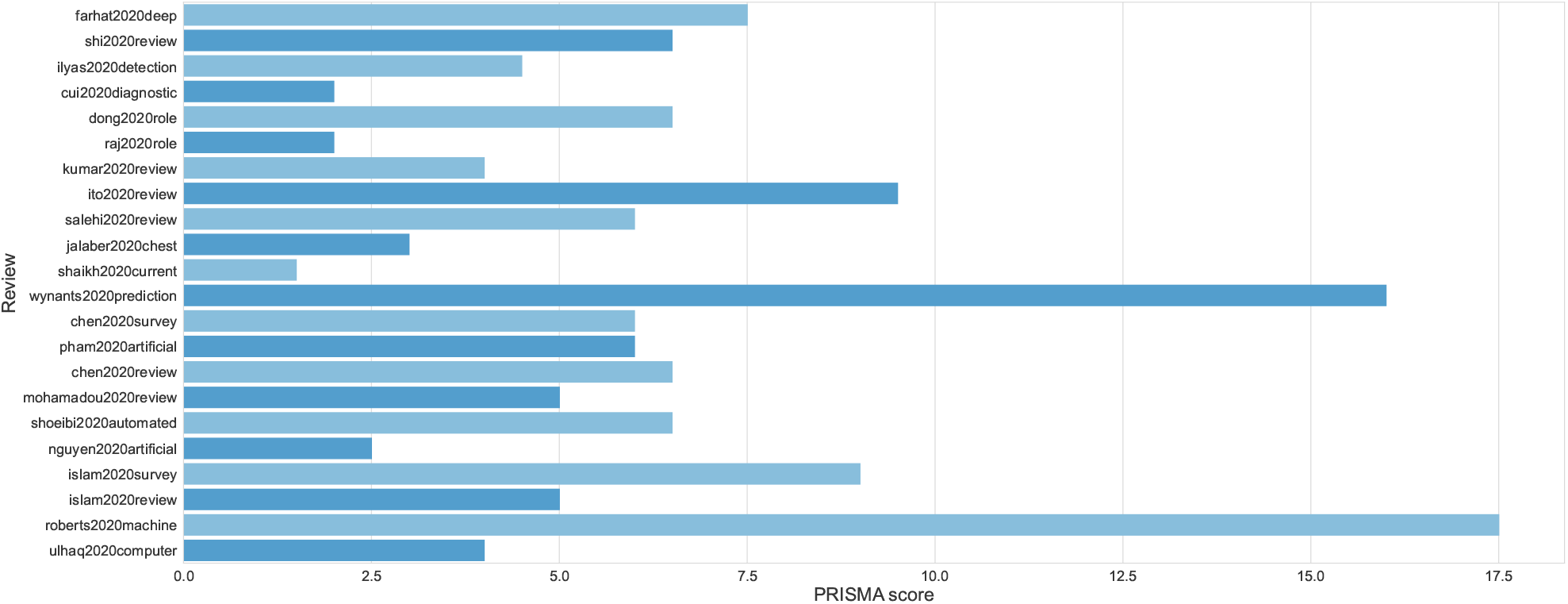
PRISMA-DTA score in each included review.

## H AMSTAR 2 heatmap

**Figure 7:**
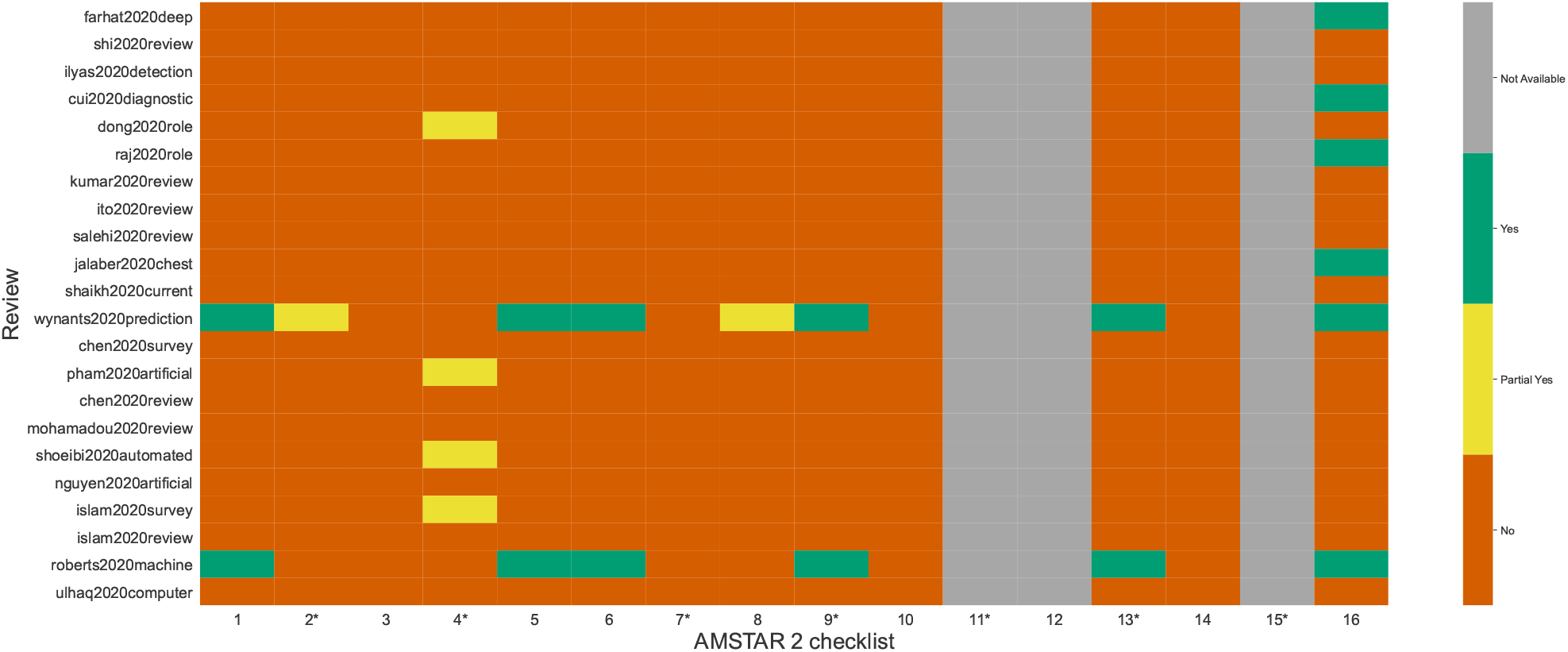
Review authors’ judgements about each AMSTAR 2 item across all included studies.

## I AMSTAR 2 results per review

**Figure 8:**
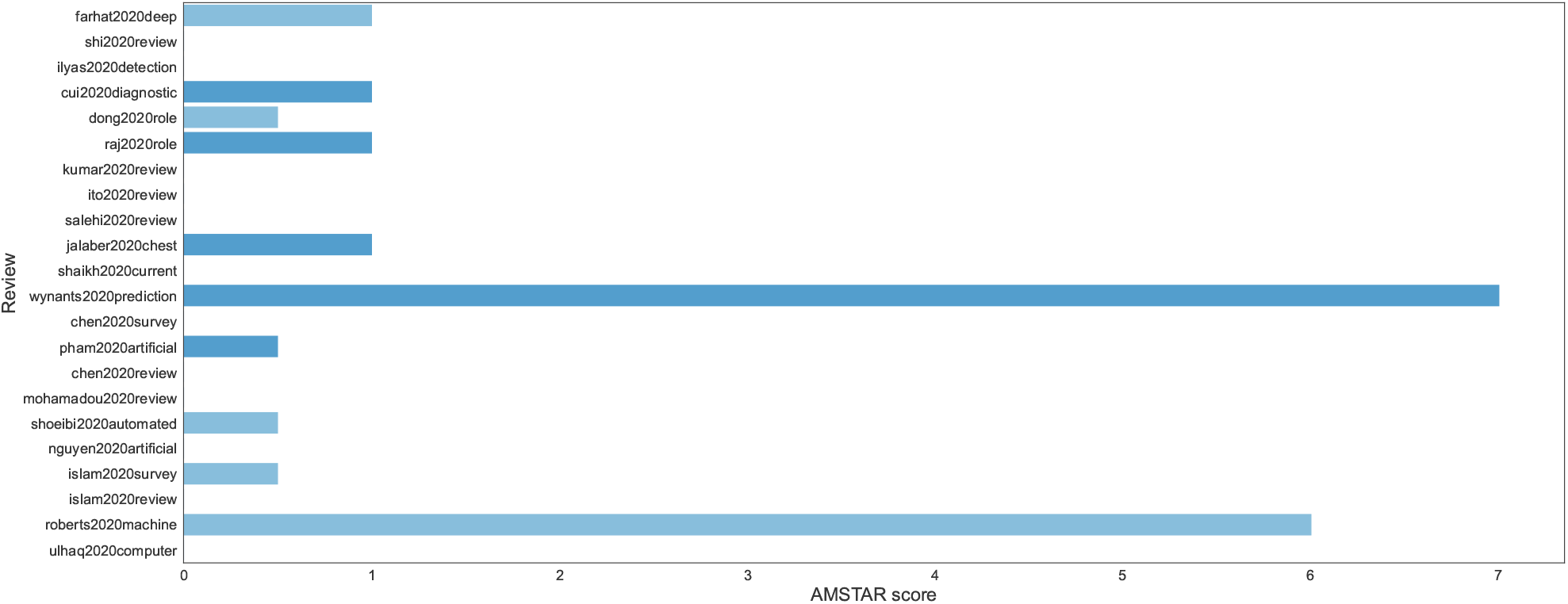
AMSTAR 2 score in each included review.

## Footnotes

1 See: https://covid19.who.int/. Accessed on May 3, 2021.

2 See: https://www.kaggle.com/allen-institute-for-ai/CORD-19-research-challenge. Accessed on May 3, 2021.

3 See: https://search.bvsalud.org/global-literature-on-novel-coronavirus-2019-ncov/. Accessed on May 3, 2021.

4 See: https://dictionary.cambridge.org/. Accessed on May 3, 2021.

5 See: https://icite.od.nih.gov/covid19/search/. Accessed on May 3, 2021.

## References

[1] Pawewl Jemiowlo, Dawid Storman, Patryk Orzechowski, and Jason Moore. Diagnosing covid-19 from medical images with artificial intelligence — an umbrella survey (registration), Nov 2020.

[2] N Kahn. New virus discovered by chinese scientists investigating pneumonia outbreak. Wall Street Journal, 2020.

[3] World Health Organization, World Health Organization, et al. Report of the who-china joint mission on coronavirus disease 2019 (covid-19), 2020.

[4] Sebastian Simon, Bernhard JH Frank, Alexander Aichmair, Philip P Manolopoulos, Martin Dominkus, Eva S Schernhammer, and Jochen G Hofstaetter. Impact of the 1st and 2nd wave of the covid-19 pandemic on primary or revision total hip and knee arthroplasty—a cross-sectional single center study. Journal of Clinical Medicine, 10(6):1260, 2021.

[5] Nasim Vahabi, Masoud Salehi, Julio D Duarte, Abolfazl Mollalo, and George Michailidis. County-level longitudinal clustering of covid-19 mortality to incidence ratio in the united states. Scientific reports, 11(1):1–22, 2021.

[6] Sho Saito, Yusuke Asai, Nobuaki Matsunaga, Kayoko Hayakawa, Mari Terada, Hiroshi Ohtsu, Shinya Tsuzuki, and Norio Ohmagari. First and second covid-19 waves in japan: A comparison of disease severity and characteristics: Comparison of the two covid-19 waves in japan. The Journal of Infection, 2020.

[7] Mario Coccia. The effects of the first and second wave of covid-19 pandemic on public health, 2020.

[8] World Health Organization et al. World health organization coronavirus disease 2019 (covid-19) situation report, 2020.

[9] Epidemiology for public health Istituto Superiore di Sanità. Covid-19 integrated surveillance: key national data, 2020. Accessed: 06 April 2021.

[10] Wei-jie Guan, Zheng-yi Ni, Yu Hu, Wen-hua Liang, Chun-quan Ou, Jian-xing He, Lei Liu, Hong Shan, Chun-liang Lei, David SC Hui, et al. Clinical characteristics of coronavirus disease 2019 in china. New England journal of medicine, 382(18):1708–1720, 2020.

[11] Trang T Le, Alba Gutierrez-Sacristan, Jiyeon Son, Chuan Hong, Andrew M South, Brett K Beaulieu-Jones, Ne Hooi Will Loh, Yuan Luo, Michele Morris, Kee Yuan Ngiam, et al. Multinational prevalence of neurological phenotypes in patients hospitalized with covid-19. medRxiv, 2021.

[12] Sandra Lopez-Leon, Talia Wegman-Ostrosky, Carol Perelman, Rosalinda Sepulveda, Paulina A Rebolledo, Angelica Cuapio, and Sonia Villapol. More than 50 long-term effects of covid-19: a systematic review and meta-analysis. Available at SSRN 3769978, 2021.

[13] Trisha Greenhalgh, Matthew Knight, Maria Buxton, Laiba Husain, et al. Management of post-acute covid-19 in primary care. Bmj, 370, 2020.

[14] Qin Sun, Haibo Qiu, Mao Huang, and Yi Yang. Lower mortality of covid-19 by early recognition and intervention: experience from jiangsu province. Annals of intensive care, 10(1):1–4, 2020.

[15] Nayaar Islam, Jean-Paul Salameh, Mariska Mg Leeflang, Lotty Hooft, Trevor A McGrath, Christian B Pol, Robert A Frank, Sakib Kazi, Ross Prager, Samanjit S Hare, et al. Thoracic imaging tests for the diagnosis of covid-19, 2020.

[16] Centers for Disease Control and Prevention. Interim guidelines for collecting, handling, and testing clinical specimens from persons for coronavirus disease 2019 (covid-19), 2020. Accessed: 06 April 2021.

[17] Wenling Wang, Yanli Xu, Ruqin Gao, Roujian Lu, Kai Han, Guizhen Wu, and Wenjie Tan. Detection of sars-cov-2 in different types of clinical specimens. Jama, 323(18):1843–1844, 2020.

[18] Jonathan J Deeks, Jacqueline Dinnes, Yemisi Takwoingi, Clare Davenport, René Spijker, Sian Taylor-Phillips, Ada Adriano, Sophie Beese, Janine Dretzke, Lavinia Ferrante di Ruffano, et al. Antibody tests for identification of current and past infection with sars-cov-2. Cochrane Database of Systematic Reviews, 2020.

[19] David L Smith, John-Paul Grenier, Catherine Batte, and Bradley Spieler. A characteristic chest radiographic pattern in the setting of covid-19 pandemic. Radiology: Cardiothoracic Imaging, 2(5):e200280, 2020.

[20] Joanne Cleverley, James Piper, and Melvyn M Jones. The role of chest radiography in confirming covid-19 pneumonia. bmj, 370, 2020.

[21] Felix Chua, Darius Armstrong-James, Sujal R Desai, Joseph Barnett, Vasileios Kouranos, Onn Min Kon, Ricardo José, Rama Vancheeswaran, Michael R Loebinger, Joyce Wong, et al. The role of ct in case ascertainment and management of covid-19 pneumonia in the uk: insights from high-incidence regions. The Lancet Respiratory Medicine, 8(5):438–440, 2020.

[22] Tao Ai, Zhenlu Yang, Hongyan Hou, Chenao Zhan, Chong Chen, Wenzhi Lv, Qian Tao, Ziyong Sun, and Liming Xia. Correlation of chest ct and rt-pcr testing for coronavirus disease 2019 (covid-19) in china: a report of 1014 cases. Radiology, 296(2):E32–E40, 2020.

[23] Jae Young Park, Rosemary Freer, Richard Stevens, Soneji. Neil, and Nicholas Jones. The accuracy of chest ct in the diagnosis of covid-19: An umbrella review.

[24] Travers Ching, Daniel S Himmelstein, Brett K Beaulieu-Jones, Alexandr A Kalinin, Brian T Do, Gregory P Way, Enrico Ferrero, Paul-Michael Agapow, Michael Zietz, Michael M Hoffman, et al. Opportunities and obstacles for deep learning in biology and medicine. Journal of The Royal Society Interface, 15(141):20170387, 2018.

[25] Ahmed Hosny, Chintan Parmar, John Quackenbush, Lawrence H Schwartz, and Hugo JWL Aerts. Artificial intelligence in radiology. Nature Reviews Cancer, 18(8):500–510, 2018.

[26] Morgan P. McBee, Omer A. Awan, Andrew T. Colucci, Comeron W. Ghobadi, Nadja Kadom, Akash P. Kansagra, Srini Tridandapani, and William F. Auffermann. Deep learning in radiology. Academic Radiology, 25(11):1472–1480, 2018.

[27] Pu Wang, Tyler M Berzin, Jeremy Romek Glissen Brown, Shishira Bharadwaj, Aymeric Becq, Xun Xiao, Peixi Liu, Liangping Li, Yan Song, D. Zhang, et al. Real-time automatic detection system increases colonoscopic polyp and adenoma detection rates: a prospective randomised controlled study. Gut, 68(10):1813–1819, 2019.

[28] Xiaoxuan Liu, Livia Faes, Aditya U Kale, Siegfried K Wagner, Dun Jack Fu, Alice Bruynseels, Thushika Mahendiran, Gabriella Moraes, Mohith Shamdas, Christoph Kern, et al. A comparison of deep learning performance against health-care professionals in detecting diseases from medical imaging: a systematic review and meta-analysis. The lancet digital health, 1(6):e271–e297, 2019.

[29] Jessica Loo, Traci E Clemons, Emily Y Chew, Martin Friedlander, Glenn J Jaffe, and Sina Farsiu. Beyond performance metrics: automatic deep learning retinal oct analysis reproduces clinical trial outcome. Ophthalmology, 127(6):793–801, 2020.

[30] Joseph Bullock, Alexandra Luccioni, Katherine Hoffman Pham, Cynthia Sin Nga Lam, and Miguel Luengo-Oroz. Mapping the landscape of artificial intelligence applications against covid-19. Journal of Artificial Intelligence Research, 69:807–845, 2020.

[31] Yanfei Li, Liujiao Cao, Ziyao Zhang, Liangying Hou, Yu Qin, Xu Hui, Jing Li, Haitong Zhao, Gecheng Cui, Xudong Cui, et al. Reporting and methodological quality of covid-19 systematic reviews needs to be improved: an evidence mapping. Journal of Clinical Epidemiology, 2021.

[32] Wim Naudé. Artificial intelligence vs covid-19: limitations, constraints and pitfalls. AI & society, 35(3):761–765, 2020.

[33] Terence J Quinn, Jennifer K Burton, Ben Carter, Nicola Cooper, Kerry Dwan, Ryan Field, Suzanne C Freeman, Claudia Geue, Ping-Hsuan Hsieh, Kris McGill, et al. Following the science? comparison of methodological and reporting quality of covid-19 and other research from the first wave of the pandemic. BMC medicine, 19(1):1–10, 2021.

[34] Coronaviridae Study Group of the International et al. The species severe acute respiratory syndrome-related coronavirus: classifying 2019-ncov and naming it sars-cov-2. Nature microbiology, 5(4):536, 2020.

[35] Pawewl Jemiowlo, Dawid Storman, Jason H Moore, and Patryk Orzechowski. Diagnosing covid-19 from medical images with artificial intelligence – an umbrella survey, Nov 2020.

[36] BJ Copeland. Artificial intelligence: Definition, examples, and applications. Encyclopedia Britannica, 2020.

[37] Lisheng Wang, Yiru Wang, Dawei Ye, and Qingquan Liu. Review of the 2019 novel coronavirus (sars-cov-2) based on current evidence. International journal of antimicrobial agents, 55(6):105948, 2020.

[38] Cornelius T Leondes. Medical Imaging Systems Techniques and Applications: Computational Techniques, volume 6. CRC Press, 1998.

[39] KC Santosh, Sameer Antani, Devanur S Guru, and Nilanjan Dey. Medical Imaging: Artificial Intelligence, Image Recognition, and Machine Learning Techniques. CRC Press, 2019.

[40] Alexei Botchkarev. Performance metrics (error measures) in machine learning regression, forecasting and prognostics: Properties and typology. arXiv preprint 1809.03006, 2018.

[41] Mourad Ouzzani, Hossam Hammady, Zbys Fedorowicz, and Ahmed Elmagarmid. Rayyan—a web and mobile app for systematic reviews. Systematic reviews, 5(1):210, 2016.

[42] Pawewl Jemiowlo and Dawid Storman. Quality assessment of systematic reviews (qasr), Jun 2020.

[43] Beverley J Shea, Barnaby C Reeves, George Wells, Micere Thuku, Candyce Hamel, Julian Moran, David Moher, Peter Tugwell, Vivian Welch, Elizabeth Kristjansson, et al. Amstar 2: a critical appraisal tool for systematic reviews that include randomised or non-randomised studies of healthcare interventions, or both. bmj, 358:j4008, 2017.

[44] Matthew DF McInnes, David Moher, Brett D Thombs, Trevor A McGrath, Patrick M Bossuyt, Tammy Clifford, Jeremie F Cohen, Jonathan J Deeks, Constantine Gatsonis, Lotty Hooft, et al. Preferred reporting items for a systematic review and meta-analysis of diagnostic test accuracy studies: the prisma-dta statement. Jama, 319(4):388–396, 2018.

[45] Jin-long Li, Long Ge, Ji-chun Ma, Qiao-ling Zeng, Lu Yao, Ni An, Jie-xian Ding, Yu-hong Gan, and Jin-hui Tian. Quality of reporting of systematic reviews published in “evidence-based” chinese journals. Systematic reviews, 3(1):1–6, 2014.

[46] Feng Shi, Jun Wang, Jun Shi, Ziyan Wu, Qian Wang, Zhenyu Tang, Kelei He, Yinghuan Shi, and Dinggang Shen. Review of artificial intelligence techniques in imaging data acquisition, segmentation and diagnosis for covid-19. IEEE reviews in biomedical engineering, 4 2020.

[47] Muhammad Ilyas, Hina Rehman, and Amine Näit-Ali. Detection of covid-19 from chest x-ray images using artificial intelligence: An early review. arXiv preprint 2004.05436v1, 4 2020.

[48] Di Dong, Zhenchao Tang, Shuo Wang, Hui Hui, Lixin Gong, Yao Lu, Zhong Xue, Hongen Liao, Fang Chen, Fan Yang, et al. The role of imaging in the detection and management of covid-19: a review. IEEE reviews in biomedical engineering, 4 2020.

[49] Rintaro Ito, Shingo Iwano, and Shinji Naganawa. A review on the use of artificial intelligence for medical imaging of the lungs of patients with coronavirus disease 2019. Diagnostic and Interventional Radiology, 26(5):443, 5 2020.

[50] Aishwarya Kumar, Puneet Kumar Gupta, and Ankita Srivastava. A review of modern technologies for tackling covid-19 pandemic. Diabetes & Metabolic Syndrome: Clinical Research & Reviews, 14(4):569–573, 5 2020.

[51] Vimal Raj. Role of chest radiograph (cxr) in covid-19 diagnosis and management. Journal Of The Indian Medical Association, 118(5):14–19, 5 2020.

[52] Feiyun Cui and H Susan Zhou. Diagnostic methods and potential portable biosensors for coronavirus disease 2019. Biosensors and Bioelectronics, 165:112349, 6 2020.

[53] Carole Jalaber, Thibaut Lapotre, Thibaud Morcet-Delattre, Félix Ribet, Stéphane Jouneau, and Mathieu Lederlin. Chest ct in covid-19 pneumonia: A review of current knowledge. Diagnostic and Interventional Imaging, 101(7-8):431–437, 6 2020.

[54] Ahmad Waleed Salehi, Preety Baglat, and Gaurav Gupta. Review on machine and deep learning models for the detection and prediction of coronavirus. Materials Today: Proceedings, 33:3896–3901, 6 2020.

[55] Hanan Farhat, George E Sakr, and Rima Kilany. Deep learning applications in pulmonary medical imaging: recent updates and insights on covid-19. Machine vision and applications, 31(6):1–42, 7 2020.

[56] Faiq Shaikh, Michael Anderson, M Rizwan Sohail, Francisca Mulero, Omer Awan, Diana Dupont-Roettger, Olga Kubassova, Jamshid Dehmsehki, and Sotirios Bisdas. Current landscape of imaging and the potential role for artificial intelligence in the management of covid-19. Current Problems in Diagnostic Radiology, 6 2020.

[57] Laure Wynants, Ben Van Calster, Gary S Collins, Richard D Riley, Georg Heinze, Ewoud Schuit, Marc MJ Bonten, Darren L Dahly, Johanna AA Damen, Thomas PA Debray, et al. Prediction models for diagnosis and prognosis of covid-19: systematic review and critical appraisal. bmj, 369, 7 2020.

[58] Jianguo Chen, Kenli Li, Zhaolei Zhang, Keqin Li, and Philip S Yu. A survey on applications of artificial intelligence in fighting against covid-19. arXiv preprint 2007.02202v1, 7 2020.

[59] Quoc-Viet Pham, Dinh C Nguyen, Thien Huynh-The, Won-Joo Hwang, and Pubudu N Pathirana. Artificial intelligence (ai) and big data for coronavirus (covid-19) pandemic: A survey on the state-of-the-arts. IEEE Access, 8:130820–130839, 7 2020.

[60] Delong Chen, Shunhui Ji, Fan Liu, Zewen Li, and Xinyu Zhou. A review of automated diagnosis of covid-19 based on scanning images, 7 2020.

[61] Youssoufa Mohamadou, Aminou Halidou, and Pascalin Tiam Kapen. A review of mathematical modeling, artificial intelligence and datasets used in the study, prediction and management of covid-19. Applied Intelligence, 50(11):3913–3925, 7 2020.

[62] Afshin Shoeibi, Marjane Khodatars, Roohallah Alizadehsani, Navid Ghassemi, Mahboobeh Jafari, Parisa Moridian, Ali Khadem, Delaram Sadeghi, Sadiq Hussain, Assef Zare, et al. Automated detection and forecasting of covid-19 using deep learning techniques: A review. arXiv preprint 2007.10785v3, 7 2020.

[63] Thanh Thi Nguyen. Artificial intelligence in the battle against coronavirus (covid-19): a survey and future research directions. arXiv preprint 2008.07343v1, 7 2020.

[64] Muhammad Nazrul Islam, Toki Tahmid Inan, Suzzana Rafi, Syeda Sabrina Akter, Iqbal H Sarker, and AKM Islam. A survey on the use of ai and ml for fighting the covid-19 pandemic. arXiv preprint 2008.07449v1, 8 2020.

[65] Md. Milon Islam, Fakhri Karray, Reda Alhajj, and Jia Zeng. A review on deep learning techniques for the diagnosis of novel coronavirus (covid-19), 8 2020.

[66] Michael Roberts, Derek Driggs, Matthew Thorpe, Julian Gilbey, Michael Yeung, Stephan Ursprung, Angelica I Aviles-Rivero, Christian Etmann, Cathal McCague, Lucian Beer, et al. Machine learning for covid-19 detection and prognostication using chest radiographs and ct scans: a systematic methodological review. arXiv preprint 2008.06388v1, 8 2020.

[67] Anwaar Ulhaq, Jannis Born, Asim Khan, Douglas Pinto Sampaio Gomes, Subrata Chakraborty, and Manoranjan Paul. Covid-19 control by computer vision approaches: A survey. IEEE Access, 8:179437–179456, 10 2020.

[68] Matthew J Page, David Moher, Patrick M Bossuyt, Isabelle Boutron, Tammy C Hoffmann, Cynthia D Mulrow, Larissa Shamseer, Jennifer M Tetzlaff, Elie A Akl, Sue E Brennan, et al. Prisma 2020 explanation and elaboration: updated guidance and exemplars for reporting systematic reviews. bmj, 372, 2021.

[69] Qingyu Chen, Alexis Allot, and Zhiyong Lu. Keep up with the latest coronavirus research. Nature, 579(7798):193–193, 2020.

[70] Paul P Glasziou, Sharon Sanders, and Tammy Hoffmann. Waste in covid-19 research, 2020.

[71] Alex John London and Jonathan Kimmelman. Against pandemic research exceptionalism. Science, 368(6490):476–477, 2020.

[72] Elisabeth Mahase. Covid-19: 146 researchers raise concerns over chloroquine study that halted who trial. BMJ, 369, 2020.

[73] John PA Ioannidis. Coronavirus disease 2019: the harms of exaggerated information and non-evidencebased measures, 2020.

[74] Marko Zdravkovic, Joana Berger-Estilita, Bogdan Zdravkovic, and David Berger. Scientific quality of covid-19 and sars cov-2 publications in the highest impact medical journals during the early phase of the pandemic: A case control study. PloS one, 15(11):e0241826, 2020.

[75] Richard G Jung, Pietro Di Santo, Cole Clifford, Graeme Prosperi-Porta, Stephanie Skanes, Annie Hung, Simon Parlow, Sarah Visintini, F Daniel Ramirez, Trevor Simard, et al. Methodological quality of covid-19 clinical research. Nature communications, 12(1):1–10, 2021.

[76] Yang Yu, Qianling Shi, Peng Zheng, Lei Gao, Haiyuan Li, Pengxian Tao, Baohong Gu, Dengfeng Wang, and Hao Chen. Assessment of the quality of systematic reviews on covid-19: A comparative study of previous coronavirus outbreaks. Journal of medical virology, 92(7):883–890, 2020.

[77] David Moher, Alessandro Liberati, JAD Tetzlaff, and Douglas G Altman. Prisma 2009 flow diagram.The PRISMA statement, 6:97, 2009.

[78] Myura Nagendran, Yang Chen, Christopher A Lovejoy, Anthony C Gordon, Matthieu Komorowski, Hugh Harvey, Eric J Topol, John PA Ioannidis, Gary S Collins, and Mahiben Maruthappu. Artificial intelligence versus clinicians: systematic review of design, reporting standards, and claims of deep learning studies. bmj, 368, 2020.

[79] Monika Storman, Dawid Storman, Katarzyna W Jasinska, Mateusz J Swierz, and Malgorzata M Bala. The quality of systematic reviews/meta-analyses published in the field of bariatrics: A cross-sectional systematic survey using amstar 2 and robis. Obesity Reviews, 21(5):e12994, 2020.

[80] Victoria Leclercq, Charlotte Beaudart, Ezio Tirelli, and Olivier Bruyère. Psychometric measurements of amstar 2 in a sample of meta-analyses indexed in psycinfo. Journal of clinical epidemiology, 119:144– 145, 2020.

[81] Dawid Pieper, Robert C Lorenz, Tanja Rombey, Anja Jacobs, Olesja Rissling, Simone Freitag, and Katja Matthias. Authors should clearly report how they derived the overall rating when applying amstar 2—a cross-sectional study. Journal of Clinical Epidemiology, 129:97–103, 2021.

[82] ESHRE Capri Workshop Group. Protect us from poor-quality medical research. Human Reproduction, 33(5):770–776, 2018.

[83] International Committee of Medical Journal Editors et al. Uniform requirements for manuscripts submitted to biomedical journals: Writing and editing for biomedical publication international committee of medical journal editors updated october 2005 (www.icmje.org.). Indian Journal of Pharmacology, 38(2):149, 2006.

[84] Claire Johnson. Repetitive, duplicate, and redundant publications: a review for authors and readers. Journal of manipulative and physiological therapeutics, 29(7):505–509, 2006.

[85] Veronica Yank and Deborah Barnes. Consensus and contention regarding redundant publications in clinical research: cross-sectional survey of editors and authors. Journal of Medical Ethics, 29(2):109–114, 2003.

[86] Edward J Huth. Repetitive and divided publication. Ethical issues in biomedical publication, pages 112–136, 2000.

[87] Edward M Corrado. The importance of open access, open source, and open standards for libraries, 2005.

[88] Brett K Beaulieu-Jones and Casey S Greene. Reproducibility of computational workflows is automated using continuous analysis. Nature biotechnology, 35(4):342–346, 2017.

[89] Andrew AS Soltan, Samaneh Kouchaki, Tingting Zhu, Dani Kiyasseh, Thomas Taylor, Zaamin B Hussain, Tim Peto, Andrew J Brent, David W Eyre, and David A Clifton. Rapid triage for covid-19 using routine clinical data for patients attending hospital: development and prospective validation of an artificial intelligence screening test. The Lancet Digital Health, 3(2):e78–e87, 2021.

[90] The Lancet Digital Health. Artificial intelligence for covid-19: saviour or saboteur? The Lancet. Digital Health, 3(1):e1, 2021.

[91] John Mongan, Linda Moy, and Charles E. Kahn. Checklist for artificial intelligence in medical imaging (claim): A guide for authors and reviewers. Radiology: Artificial Intelligence, 2(2):e200029, 2020.

[92] Debora Gil, Katerine Diaz-Chito, Carles Sanchez, and Aura Hernandez-Sabate. Early screening of sars-cov-2 by intelligent analysis of x-ray images. arXiv preprint 2005.13928, 2020.

[93] AS Albahri, Rula A Hamid, Jwan K Alwan, ZT Al-Qays, AA Zaidan, BB Zaidan, AOS Albahri, AH AlAmoodi, Jamal Mawlood Khlaf, EM Almahdi, et al. Role of biological data mining and machine learning techniques in detecting and diagnosing the novel coronavirus (covid-19): a systematic review. Journal of medical systems, 44:1–11, 2020.

[94] Keno K Bressem, Lisa C Adams, Christoph Erxleben, Bernd Hamm, Stefan M Niehues, and Janis L Vahldiek. Comparing different deep learning architectures for classification of chest radiographs. Scientific reports, 10(1):1–16, 2020.

[95] Vinay Chamola, Vikas Hassija, Vatsal Gupta, and Mohsen Guizani. A comprehensive review of the covid-19 pandemic and the role of iot, drones, ai, blockchain, and 5g in managing its impact. Ieee access, 8:90225–90265, 2020.

[96] Nicola Luigi Bragazzi, Haijiang Dai, Giovanni Damiani, Masoud Behzadifar, Mariano Martini, and Jianhong Wu. How big data and artificial intelligence can help better manage the covid-19 pandemic. International journal of environmental research and public health, 17(9):3176, 2020.

[97] Prashant Nagpal, Sabarish Narayanasamy, Aditi Vidholia, Junfeng Guo, Kyung Min Shin, Chang Hyun Lee, and Eric A Hoffman. Imaging of covid-19 pneumonia: Patterns, pathogenesis, and advances. The British journal of radiology, 93(1113):20200538, 2020.

[98] Agam Bansal, Rana Prathap Padappayil, Chandan Garg, Anjali Singal, Mohak Gupta, and Allan Klein. Utility of artificial intelligence amidst the covid 19 pandemic: a review. Journal of Medical Systems, 44(9):1–6, 2020.

[99] Mahdi Rezaei and Mahsa Shahidi. Zero-shot learning and its applications from autonomous vehicles to covid-19 diagnosis: A review. Intelligence-based medicine, page 100005, 2020.

[100] Amit Kharat, Vinay Duddalwar, Krishna Saoji, Ashrika Gaikwad, Viraj Kulkarni, Gunjan Naik, Rohit Lokwani, Swaraj Kasliwal, Sudeep Kondal, Tanveer Gupte, et al. Role of edge device and cloud machine learning in point-of-care solutions using imaging diagnostics for population screening. arXiv preprint 2006.13808, 2020.

[101] OS Albahri, AA Zaidan, AS Albahri, BB Zaidan, Karrar Hameed Abdulkareem, ZT Al-Qaysi, AH Alamoodi, AM Aleesa, MA Chyad, RM Alesa, et al. Systematic review of artificial intelligence techniques in the detection and classification of covid-19 medical images in terms of evaluation and benchmarking: Taxonomy analysis, challenges, future solutions and methodological aspects. Journal of infection and public health, 2020.

[102] S Manigandan, Ming-Tsang Wu, Vinoth Kumar Ponnusamy, Vinay B Raghavendra, Arivalagan Pugazhendhi, and Kathirvel Brindhadevi. A systematic review on recent trends in transmission, diagnosis, prevention and imaging features of covid-19. Process Biochemistry, 2020.

